# Socio-environmental typologies of dengue risk at multiple spatial scales of the urban landscape

**DOI:** 10.64898/2026.03.24.26349225

**Authors:** Pallavi Kache, Maria Diuk-Wasser, Mauricio Santos-Vega

## Abstract

Urbanization in the 21st century has given rise to complex socio-environmental landscapes that contribute to spatial inequalities in health, particularly in the context of emerging infectious diseases, such as dengue fever. This study employs an urban systems framework to explore the multi-dimensional drivers of dengue risk in Ibagué, Colombia, where Aedes mosquitoes thrive in diverse urban environments. We characterized the biophysical, socio-economic, and institutional properties of the urban landscape and employed hierarchical cluster analysis to define urban typologies at both census block and urban section levels. Our findings reveal significant differences in dengue incidence across these typologies, with higher rates associated with areas of high population density and commercial activity. Additionally, we examined the landscape configuration and its role in shaping dengue risk, identifying that diversity and intermixing of typologies had protective effects against dengue incidence. This research underscores the importance of considering multi-scale, socio-ecological factors in dengue risk assessments and highlights the need for targeted public health interventions that address the complex interactions within urban landscapes.

## 1. Introduction

Twenty-first-century urbanization is occurring with complex growth dynamics and unprecedented speed, contributing to highly heterogeneous landscapes (Seto et al., 2010). For many cities, rapid land use and demographic change contributes to strained government resources, inadequate urban planning, and widening social and infrastructural disparities, driving spatial inequalities in human health and well-being—including the burden of emerging infectious diseases (Deng et al., 2009; Desa, 2018b; Fragkias et al., 2013; Pandey et al., 2022; Thapa and Murayama, 2009). Among the most pressing infectious disease threats that cities face is dengue fever, transmitted by the highly urban-adapted mosquitoes, *Aedes aegypti* and *Ae. albopictus*. These species are biologically linked to humans as bloodmeal hosts and to the built environment for larval habitat. Therefore, variation in biophysical and human social conditions across cities affects both *Aedes* spp. vector ecology and pathogen transmission dynamics. The complex social, ecological, and technological processes occurring within urbanizing landscapes necessitate new approaches to conceptualizing and monitoring *Aedes*-borne disease risk within cities, particularly to inform spatially targeted dengue interventions (McPhearson et al., 2016).

Currently, the links between specific urban inequalities and dengue risk are studied separately. For example, is household income significantly linked to dengue incidence, or does higher neighborhood vegetation have a positive or negative effect on *Aedes spp*. abundance? Instead, socio-ecological factors are more likely to interact in additive, antagonistic, or synergistic ways, influencing the emergence of patterns of dengue risk (Alberti and Wang, 2022; Roy Chowdhury et al., 2011). A recent urban systems framework for *Aedes*-borne diseases by Kache et al. proposes a complex adaptive systems approach to understanding disease risk, wherein spatial patterns of disease incidence are the product of co-occurring, interacting processes across multiple scales of the urban system (Kache et al., 2022). Foundational to the framework is the idea that the urban landscape is comprised of independent spatial cells or “building blocks” that self-organize into homogeneous patches. Each cell is defined by multifactorial combinations of biophysical, socio-economic, institutional, and cultural properties relevant to *Aedes* spp. population growth or dengue transmission (Figure 1a). Complexity increases at higher spatial scales, accompanied by increasingly heterogeneous aggregation and divergent processes over time.

**Figure 1.**
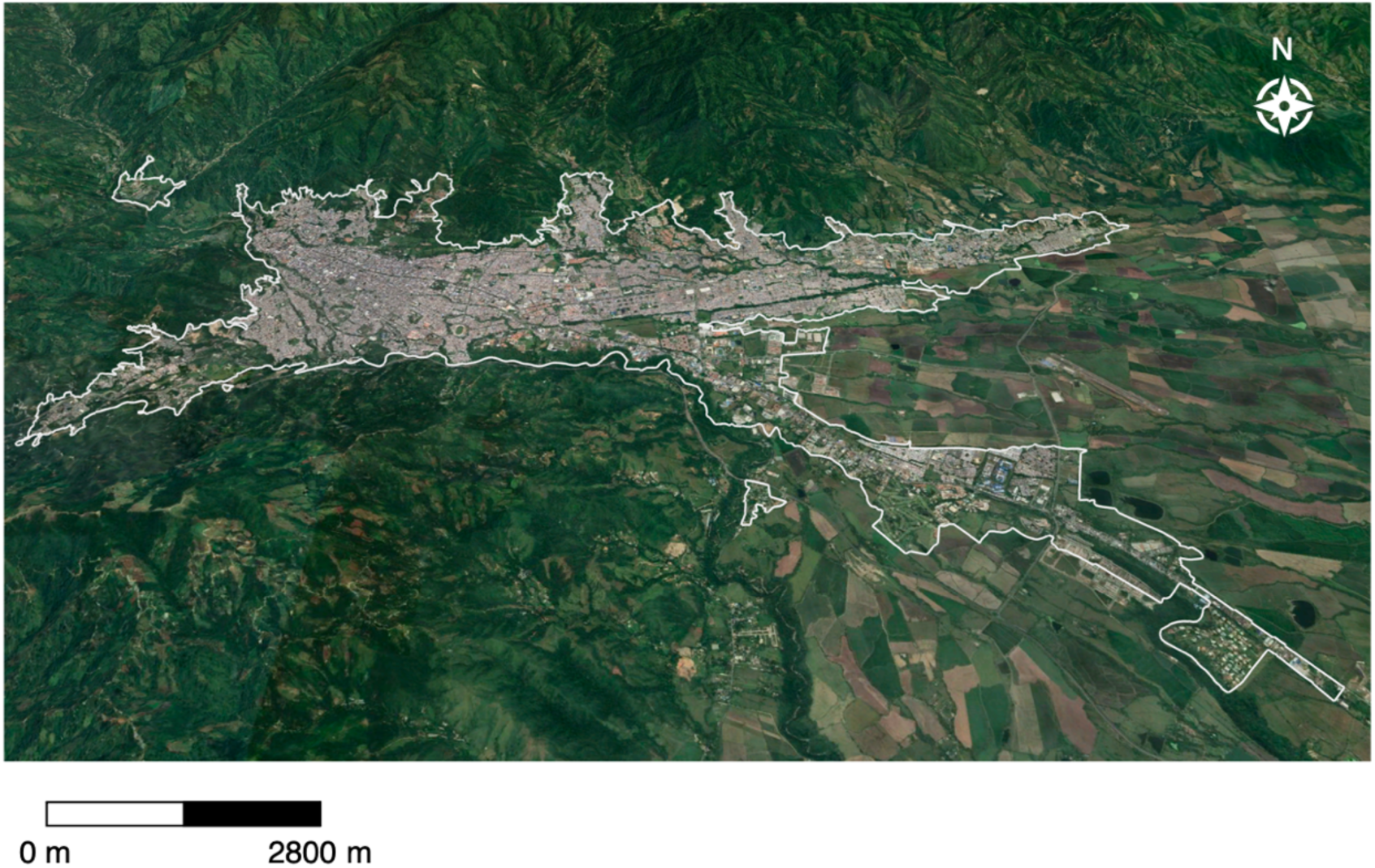
Urban extent of Ibagué, Colombia (2022) Satellite imagery of Ibagué, Colombia at a 30 cm resolution. Imagery shows land parcels of rice cultivation on the eastern border of the city, with the Andes Mountains surrounding the city on the west. The formally-recognized urban extent of Ibagué is shown with the white polygon. Imagery is from October 16, 2022, © 2023 CNES/Airbus; © 2023 Maxar Technologies, accessed via Google Earth.

While landscape composition establishes localized levels of disease risk, landscape configuration determines the patterns of risk that emerge at patch and landscape levels (Kache et al., 2022). Landscape configuration describes the spatial layout and structure of the built environment (Kropf, 2009). Patches of varying composition are arranged in unique ways, and patterns and processes emerge that have cascading impacts on *Aedes*-borne disease risk. Aggregated cells with varied building densities and impervious land cover have consequences for mosquito dispersal (Dimoudi et al., 2013; Harrington et al., 2005; Hemme et al., 2010; Honório et al., 2003; Middel et al., 2014). Additionally, depending on cell-level configuration, human populations are spatially dispersed at different densities (Romeo-Aznar et al., 2021; Seto et al., 2014).

The complex associations between dengue virus (DENV), *Aedes* spp., and human populations have prompted researchers to ask the question—what is the most appropriate geographic scale to measure dengue risk?—as the parameters and processes at one scale may be more or less influential on the ecological or epidemiological outcome at another scale (Marti et al., 2020; Scott and Morrison, 2010b). Although most dengue risk factors are likely to exhibit spatial dependence, few articles explicitly evaluate the multi-scale spatial relationships between the built environment and dengue risk (Marti et al., 2020). Understanding which aspects of composition influence disease risk at specific scales and how high-risk areas are spatially arranged can guide scale-dependent management strategies. For example, focusing indoor mosquito spraying on certain high-risk census blocks, while broadly increasing educational efforts in low-resource, low-income areas.

This work presents an operational framework for evaluating multi-dimensional drivers of dengue risk within an urban systems approach. First, we characterized the biophysical, socio-economic, and institutional features of the urban landscape using open-source datasets. Then, using unsupervised classification techniques, we identified the basic units of the landscape, or minimal spatial units with similar dengue risk profiles (i.e., urban typologies). Finally, regarding urban typologies, we asked three key questions: Are there statistical and structural differences between typologies at various spatial scales? How are typology assignments related to the level of dengue incidence? And how do typology configurations correlate with dengue incidence?

To assess dengue-landscape associations at multiple scales, we harnessed spatially disaggregated dengue information from Ibagué, Colombia, a medium-sized city at the foothills of the Andes Mountains, with year-round dengue transmission and distinct epidemic periods (Carrasquilla et al., 2021). The city is embedded in a landscape of complex topography and heterogeneous land cover (Fig. 2). East of the city are agricultural lowlands (800–900 m), while west of the city is mountainous terrain and forested land cover (1200–1450 m) (Núñez Pita, 2021). Ibagué has had a fragmented history of urban development, with a historic city center built in a gridded Spanish-colonial form and contemporary constructions characterized by sharp contrasts between dense informal housing communities, high-income gated communities, and mid- to high-rise apartment buildings (Francel, 2017). Given these heterogeneities in Ibagué’s landscape, we employed geographic and geospatial methods to inform scale-dependent management strategies for the city, applying methodologies expandable to regions around the world.

**Figure 2.**
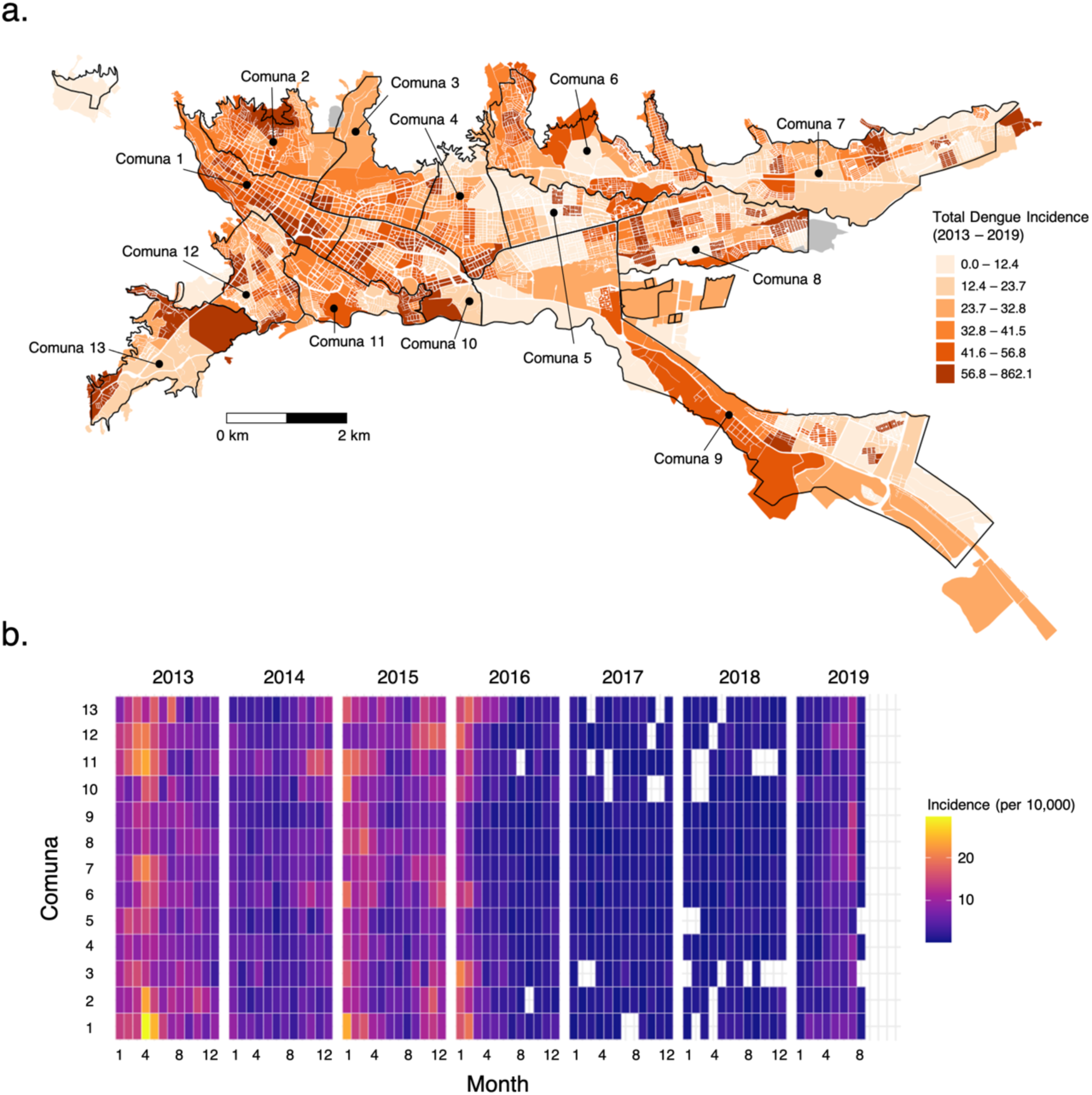
Dengue Incidence in Ibagué, Colombia (2013–2019). **a.** Total dengue incidence at the urban section level from January 2013–August 2019 per 1,000 population, shown using quantile breaks. Black outlines show the 13 *comunas* within the city. **b.** Heatmap of monthly dengue incidence per year at the *comuna* level from January 2013 – August 2019, per 10,000 population. Dengue outbreak years for Ibagué occurred in 2013, 2015, and 2019.

## 2. Methods

### Spatial units of analysis

We conducted all spatial analyses at two levels: the census block and the urban section, which are nested administrative boundaries established by the Colombian government (Figure S.1). Conceptually, these levels serve as two potential building blocks for understanding the relationship between Ibagué’s socio-environmental landscape and dengue risk. We assume that the urban section represents neighborhood-scale characteristics, and that the census block represents sub-neighborhood-scale properties. We analyzed data for 4,999 residential census blocks (with at least one reported dwelling) of the city’s 5,571 total census blocks. Residential census blocks had a median spatial extent of .2 hectares per block [Interquartile range (IQR): 0.12–0.43 hectares]. Additionally, we examined data for 269 of 271 urban sections, with a median size of 10.2 hectares (IQR: 6.6–18.3 hectares). We also looked at dengue cases at the commune level. Ibagué has 13 communes, which- unlike the census block and urban section levels- are used for administrative decision-making and are commonly recognized by residents across the city.

### Data sources

#### Dengue cases

We used dengue data collected by the Secretary of Health of Ibagué from probable and laboratory-confirmed reports during January 2013–July 2019 (Supplemental Methods). This dataset included case information, such as patients’ dates of symptom onset, dates of medical attention, and reported addresses of residence. Addresses were geolocated to longitude and latitude coordinates by a third-party georeferencing service in Colombia, Servinformacion. Records that could not be geolocated to a residential block within the city, such as those falling within public spaces (e.g., parks, public squares) or rural areas surrounding the urban boundary (i.e., veredas), were excluded from the analysis. The point data were then aggregated to the various spatial units of analysis.

#### Socio-environmental data

We compiled data on 18 candidate variables representing biophysical, socioeconomic, and institutional urban properties. We hypothesized that these factors contribute to the dengue infection hazard, exposure to infected Aedes spp. mosquitoes, or population vulnerability (Table S.1). We processed six environmental layers derived from open-source and remote-sensing datasets. First, we calculated the density of street intersections per spatial unit using OpenStreetMap street line data, focusing on major roads, including highways and primary, secondary, and tertiary roads. Next, we extracted remotely sensed variables, including vegetation (Normalized Difference Vegetation Index, 30-m resolution), built-up land cover intensity and height (built-up intensity, 30-m resolution; built-up height, 100-m resolution), elevation (30-m resolution), and slope (30-m resolution). These environmental data layers were extracted separately as shapefiles for the two analysis units, maintaining the same spatial resolution. Data extraction and manipulation were performed using Google Earth Engine, R version 4.2.1, and QGIS 3.28.1-Firenze (Supplemental Methods).

We collected data on institutional and socioeconomic characteristics from the 2018 Colombian national census (DANE, 2018) at the census block level. Institutional variables included the proportions of dwellings with access to public water, sewage systems, and trash collection. Socio-economic variables analyzed included the percentages of buildings used for services or commerce, unoccupied dwellings, population density, household occupant density, internet access, children under 10 years old, residents with primary education, and dwellings classified as Estrato 1—an indicator of low socio-economic status defined by the government to set utility prices? (Supplemental Methods). The data were provided as raw integers, enabling the aggregation of census information from the block level up to the urban section level without the need for averaging across boundaries, as census blocks are fully contained within urban sectors.

### Multi-dimensional landscape composition metric

#### Exploratory analyses

To examine how these 18 candidate variables covaried and how this relationship changed across scales, we conducted correlation analyses at each spatial unit of analysis. We performed principal component analyses (PCAs) using all combinations of low-correlation socio-environmental variables, based on the combinatorial output from the “expand.grid” function in R. From the PCA, we retained variables that contributed more than 40% to the explained variance across the first five dimensions. Analyses and visualization were conducted using the “ggcorrplot” and “FactoMine” packages in R. For the selected socio-environmental co-variates, we assessed the degree of spatial autocorrelation present at different distances in the landscape (from 0–6 km) using global Moran’s *I* statistic, conducted with the “spdep” and “rgeos” packages in R.

#### Hierarchical clustering algorithm for urban typology assignment

We conducted unsupervised hierarchical cluster analysis to group spatial units within a multi-dimensional feature space, aiming to uncover their internal structure based on selected covariates (Foot, 2017). Dissimilarity between observations was measured using Euclidean distance, while Ward’s minimum variance method assessed dissimilarity between clusters. Ward’s method minimizes total within-cluster variance at each step. The process began with an agglomerative nesting algorithm, where each observation started as a single-element cluster (Murtagh and Legendre, 2014). Then, at each step, the two most similar clusters were merged, forming larger clusters until all observations were grouped into a single cluster. The results were validated with two metrics: internal validation via the Dunn index, which evaluates cluster compactness and separation; and stability metrics including the average proportion of non-overlap (APN), average distance (AD), the average distance between means (ADM), and the figure of merit (FOM) (Supplemental Methods). The hierarchical tree was visualized as a dendrogram. This clustering assigned an urban typology to each of the 4,999 census blocks and 269 urban sections analyzed. Analyses and visualizations were performed using the “cluster”, “clValid”, and “dendextend” packages in R.

#### Validation and typology interpretation

To validate the resulting k classes, we randomly selected 10 pixels from each typology and visualized them in Google Earth (Stokes and Seto, 2019). We then interpreted and labeled the classes with designations such as “high-elevation, high density” or “El Centro commercial district,” based on socio-environmental covariates, spatial associations (i.e., core versus periphery), Google Earth imagery, and landscape characterizations utilizing the authors’ knowledge of Ibagué.

### Landscape configuration metrics

To analyze the spatial arrangement of typologies, we calculated landscape metrics based on the categorical typology assignments. We rasterized the spatial vector layer of typology data using a 1m² cell size in QGIS 3.28.1-Firenze to capture high-resolution details. Subsequently, we created buffers of 1, 1.5, and 2 km around each census block and urban section, guided by the distance bands of spatial autocorrelation from Moran’s I. For each buffer, we computed multiple landscape metrics, including those measuring diversity, complexity, shape, and aggregation. Diversity metrics such as Shannon’s index and marginal entropy were used to assess heterogeneity in typological composition within the surrounding area. Aggregation metrics such as interspersion and juxtaposition indices evaluated the spatial mixing of classes. Patch-level shape metrics, including shape index and perimeter-area fractal dimension, examined the complexity of each spatial unit’s shape—highlighting areas of formal versus informal development. These analyses used the “landscapemetrics” package in R.

### Statistical associations between dengue cases and urban landscape metrics

We used negative binomial generalized linear models (GLMs) with a “logit” link to assess the relationship between landscape composition and configuration metrics, and dengue cases during the study period, 2013–2019, using the “MASS” package in R. We first constructed univariate models of the urban typology assignments with total dengue incidence to examine whether there were statistically significant differences in dengue incidence rates across typologies, compared to a reference typology group. We then ran a univariate model for each year separately to assess inter-annual variation in dengue incidence across typologies. We evaluated various configuration measures within the spatial buffer surrounding and encompassing each building block. We used the “dredge” function in the “MuMIn” package in R to examine possible subsets of the global models and reported models within 4 AIC from the model with the lowest AIC value, accounting for 95% or greater of the AIC weight (𝑤). We estimated the variance inflation factor (VIF) of each model to ensure all variables had a VIF < 4. All models were run at the census block level and the urban section levels.

## 3. Results

### Dengue in Ibagué

During the study period in Ibagué, outbreaks of dengue were recorded in 2013, 2015 (which persisted into the first half of 2016), and 2019. These patterns align with dengue outbreaks observed at the national level in Colombia; however, case counts for Ibagué diminished significantly more rapidly during the inter-epidemic periods in 2014 and 2016 (see Figure S.2). The incidence of dengue was at its highest in the western region of the city, particularly within urban sections and census blocks in Comunas 1, 2, 11, and 12 (Figure 2). In the eastern part of the city, urban sections and census blocks in Comuna 7 experienced the highest disease burden. Spatial autocorrelation of dengue incidence was identified at the urban section level, with a Moran’s I value of 0.34 (p < 0.01), and at the census block level, with a Moran’s I value of 0.16 (p < 0.01).

### Urban section-level analyses

#### Socio-environmental landscape composition

At the urban section level, public utilities such as access to water and sewage services demonstrated a high degree of correlation (r > 0.72), as did socio-economic variables, including the proportion of residents residing in low-estrato areas and the proportion of residents possessing a primary-level formal education (r > 0.65) (see Figure S.9). Following the elimination of six of these highly correlated variables, a principal component analysis (PCA) revealed that eight out of twelve variables contributed to 56.0% of the cumulative variance within the initial five orthogonal dimensions. These variables encompassed elevation, slope, built-up intensity, the proportion of buildings with water system access, the proportion of unoccupied buildings, the proportion of buildings designated for commercial or service use, population density, and the proportion of residents living in low socio-economic areas (i.e., Estrato 1) (refer to Figure 3; Table S.3). The first two dimensions accounted for only 45% of the variance among the eight variables, indicating significant variability in socio-environmental features that limit their utility as comprehensive predictors of landscape composition. Elevation and slope exhibited significant spatial autocorrelation across the landscape extent (4–6 km), emphasizing their global spatial patterns (see Figure S.8). Land use and socio-economic features, such as built-up intensity and the proportion of buildings with commercial or service use, demonstrated spatial autocorrelation within the 1–2 km range, whereas access to water was spatiallo autocorr up to 1km, pop density up to around 2 and then remained weakly autocorrelated at larger distances.

**Figure 3.**
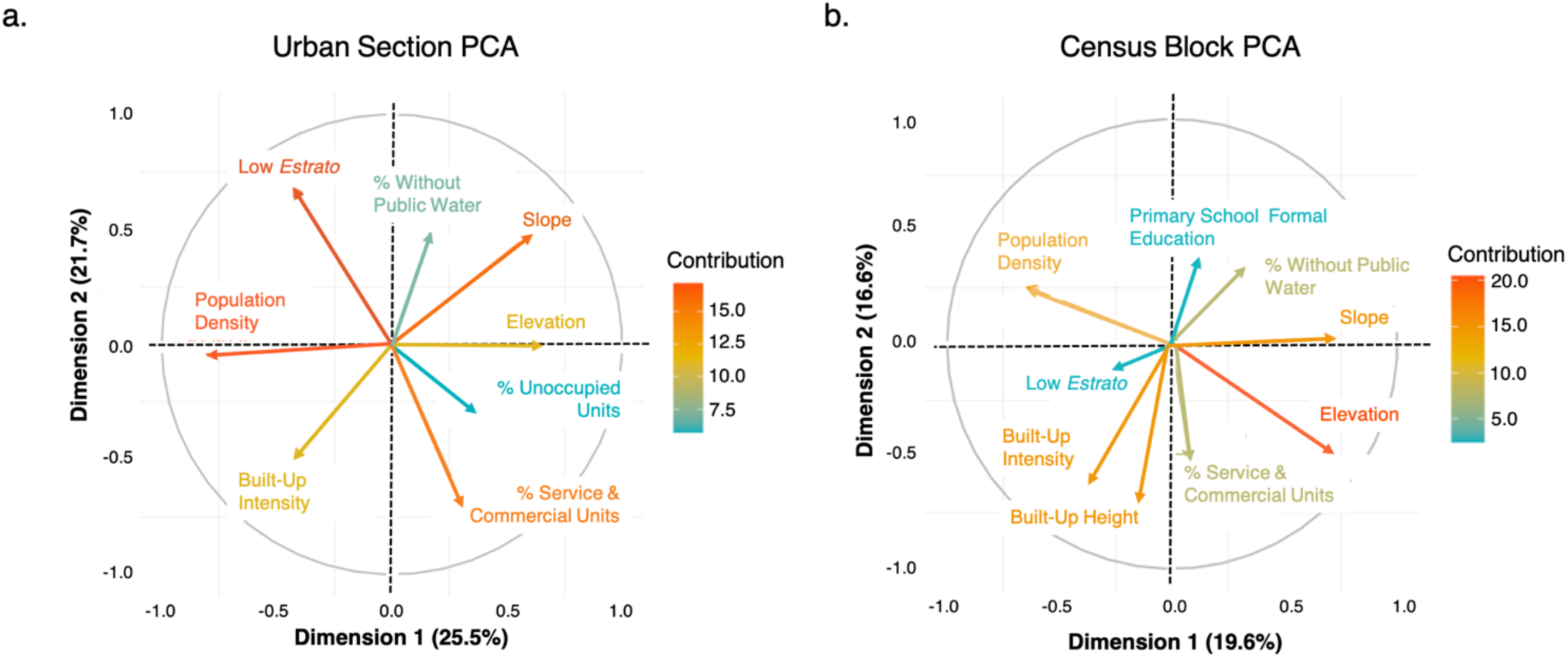
Principal component analyses (PCA) at the urban section and census block scales. **a.** For the urban section scale, the first component had an intermediate to strong positive association with elevation and slope, and a strong negative association with population density. The second component had a strong positive association with low socio-economic status (i.e., low-*estrato*) urban sections, and intermediate to strong negative associations with built-up intensity and the percent of units with service and commercial use. **b.** The first component had an intermediate to strong positive association with elevation and slope, and an intermediate negative association with population density. The second component had strong negative associations with built-up intensity and built-up height.

Hierarchical cluster analyses revealed that internal validity was highest when the urban sections were grouped into two typologies (Table S.4). These two typologies had significant differences in population density and less pronounced differences in built-up intensity, elevation, and slope. Clusters were most stable, however, when grouped into 10 typologies (using average distance and figure of merit measures) (Figure 4).

**Figure 4.**
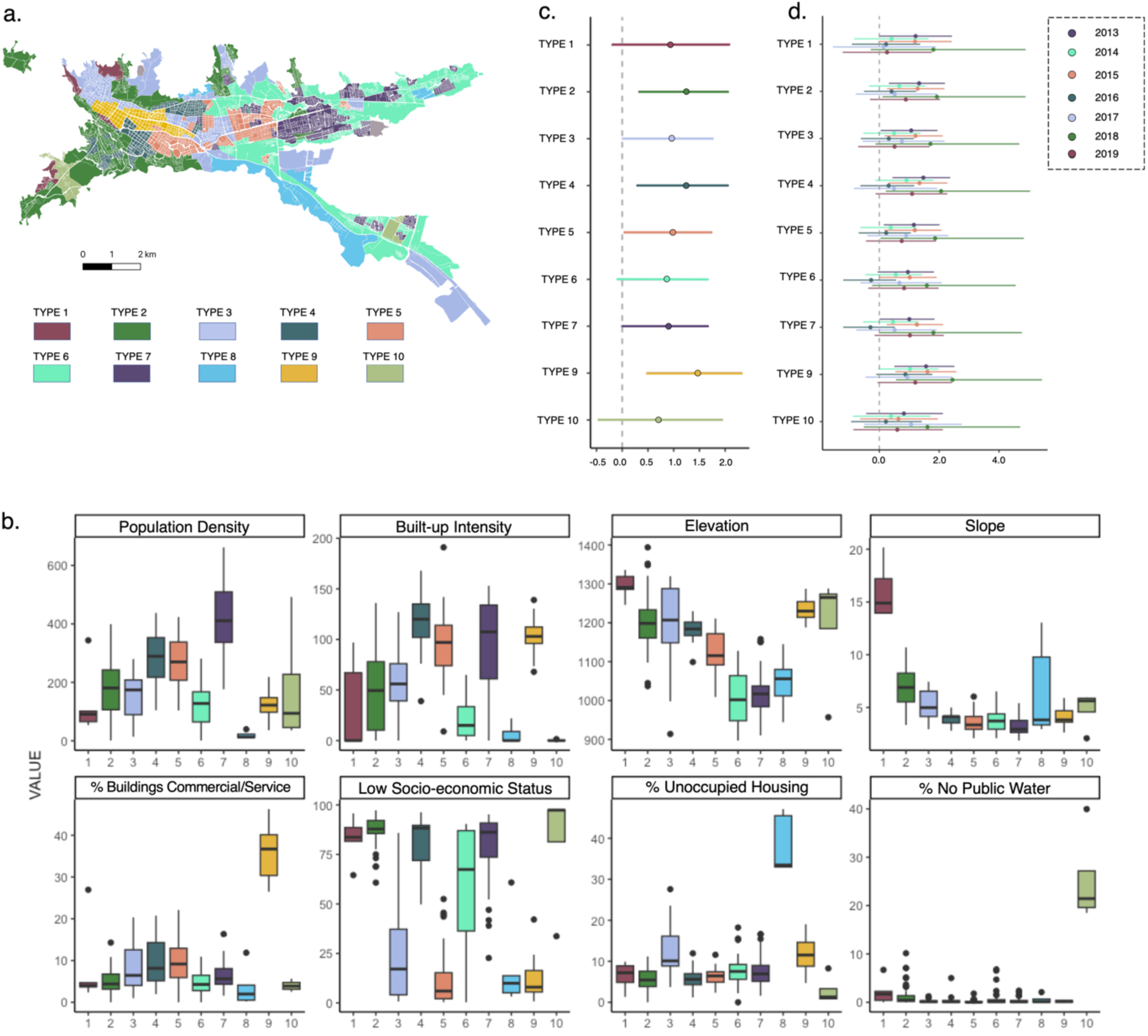
Associations between dengue incidence and landscape composition and configuration (urban section scale) **a.** The 10 socio-environmental typologies mapped onto the urban sections of Ibagué. Type 1 urban sections are characterized as high-elevation areas located in the urban periphery. Type 2 sections are characterized as being high-elevation low-socio-economic status areas (mixed urban core and periphery). Type 3 sections are characterized as high-elevation, high socio-economic status areas (mixed urban core and periphery). Type 4 sections are high-elevation high population/building density areas (located in the urban core). Type 5 are high-density mixed-use areas (located in the urban core). Type 6 are low-density areas (mixed urban core and periphery). Type 7 urban sections are high density residential areas (located in the urban core). Type 8 are low-density areas located in the urban periphery. Type 9 are high density commercial areas located in the urban core. Type 10 urban sections are low socio-economic status sections located in the urban periphery. Full characterization of the 10 urban section typologies are located in Table 3.1. **b.** Boxplots comparing the statistical values for the eight socio-environmental covariates retained in the hierarchical clustering algorithm. **c.** Coefficient plot showing the effect of nine typologies on total dengue cases during January 2013–July 2019 with 95% confidence intervals, compared to Typology 8 as a reference variable and offset by the population size of the urban section. **d.** Coefficient plot showing the effect of nine typologies on annual dengue cases during 2013–2019 with 95% confidence intervals, compared to Typology 8 as a reference variable and offset by population size of the urban section.

Typology 8 was characterized by low population density and a high proportion of unoccupied housing, and high socio-economic status (Table 3.1). Typology 9 sections were situated at high elevations, with high built-up intensity, intermediate population density, and high socio-economic status; additionally, it was the only typology with a high proportion of buildings with commercial or service uses. While Typology 4 sections were spatially adjacent to Typology 9 sections and also situated at high elevations with high built-up intensity, they had a high population density, low socio-economic status, and a low proportion of commercial or service building use. Finally, Typology 2 sections were also located at high elevations, although they were situated on the periphery of the city at a steep slope; here, urban sections had a low built-up intensity and intermediate population density, with a high proportion of residences designated as *Estrato*-1.

#### Association between socio-environmental typologies and dengue incidence

Typology 8 had the lowest rates of dengue incidence during 2013–2019, and was therefore selected as the reference group for the univariate negative binomial regression. Compared to Typology 8, dengue incidence rates were highest for Typology 9 [Incidence Rate Ratio (IRR) = 4.34, p < 0.01], Typology 2 (IRR=3.50, p < 0.01), and Typology 4 (IRR = 3.46, p < 0.01) (Table 3). Compared to the reference typology, dengue incidence rates were also significantly higher for Typology 5 (IRR=2.65, p<0.05), Typology 3 (IRR=2.62, p<0.05), and Typology 7 (IRR=2.48, p < 0.05). Although located at different elevations and with varying socio-economic profiles, all three typologies were characterized by high population densities. When examining models separately for each year during 2013 to 2019, we observed variation in the IRR for Typologies 3, 5, and 7 (Figure 4d). However, Typologies 2 and 9 remained the sections with highest IRR across years.

#### Landscape configuration metrics

Using the landscape composition model as a baseline model, we tested multiple shape, aggregation, and diversity landscape metrics at 1, 1.5, and 2 km buffers, based on the distances of significant positive spatial autocorrelation for socio-economic and built-up intensity variables indicated by Moran’s *I* analyses (Figure S.7). The best-fit models showed negative associations of dengue incidence with the Shannon’s diversity index within a 2 km buffer (IRR = 0.58, p < 0.01), and positive associations with contagion metrics (measuring the intermixing of different typologies) within a 2 km buffer (IRR=1.04, p<0.01). Shape metrics, including the perimeter-area fractal dimension and the mean fractal dimension index for each patch were not significant at any of the spatial buffer sizes tested.

### Census block analyses

#### Socio-environmental landscape composition

At the census block level, there was a high degree of correlation among public service variables (r > 0.52) as well as among socio-economic variables, in particular the proportion of the census block with internet access (r=0.66) (Figure S.9). Additionally, low socio-economic status and unoccupied housing were significantly negatively correlated (r = -0.74), indicating that unoccupied housing is primarily located in middle to high socio-economic status areas in Ibagué. After removing highly correlated variables, through principal component analyses we found that nine out of 12 variables contributed to 80.0% of the cumulative variance within the first five dimensions, including: elevation, slope, built-up intensity, built height, the proportion of buildings with water systems access, the proportion of buildings with commercial or service use, population density, the proportion living in low socio-economic areas, and the proportion with a primary-level formal education. The first two dimensions explained 38.2% of the variance contained within these nine variables, with elevation and slope (alongside population density) contributing highly to the first dimension. The second dimension was characterized by built-up intensity and height. Elevation and slope were significantly spatially autocorrelated across the spatial extent of the city (4–6 km) (Figure S.8). Land use and socio-economic features were spatially autocorrelated at intermediate to global distances (2–4 km), while public service and socio-economic variables were spatially autocorrelated at local to intermediate distances (1–2 km).

Hierarchical cluster analyses revealed that internal validity was highest when the census blcoks were grouped into two typologies. These two typologies had significant differences in population density and less pronounced differences in built-up intensity, elevation, and slope. Measures of cluster stability were highest, however, with six and 10 typologies (using average distance and figure of merit measures, respectively). We therefore regressed total dengue cases on both typology assignments using negative binomial GLMs.

#### Association between socio-environmental typologies and dengue incidence

The 10-typology structure had a lower AIC value (ΔAIC = 54) compared to the six-typology structure, indicating a better model fit, therefore we selected this as a baseline model. For the 10-typology model, Type 7 census blocks, characterized by lower built-up intensity, higher formal education, and higher socio-economic status compared to other typology classes had the lowest mean dengue incidence (2.82 cases per 1,000 people, 2013–2019) and was selected as the reference group. Compared to Type 7 census blocks, dengue risk was highest for Type 6 (IRR=2.67, p < 0.001) and Type 5 census blocks (IRR=1.97, p < 0.001), offset by the population of these blocks (Table 4, Figure 5c). These typologies consistently had a higher IRR compared to the reference group when negative binomial GLMs were run separately for each year (Figure 5d). Type 6 census blocks were spatially dispersed throughout the landscape. Counterintuitively, these blocks had higher socio-economic status, higher formal education levels, and lower population densities compared to other typologies; although were also characterized by higher proportions of non-residential, commercial lots and a higher built-up intensity compared to other typologies (Table 2). Type 5 blocks were largely aggregated within the historic and commercial district of “*El Centro*,” although the blocks also extended into the eastern periphery of the city. Similar to Type 6, Type 5 blocks had high built-up intensity and commercial use, although were of significantly lower socio-economic status compared to Type 6 blocks.

**Figure 5.**
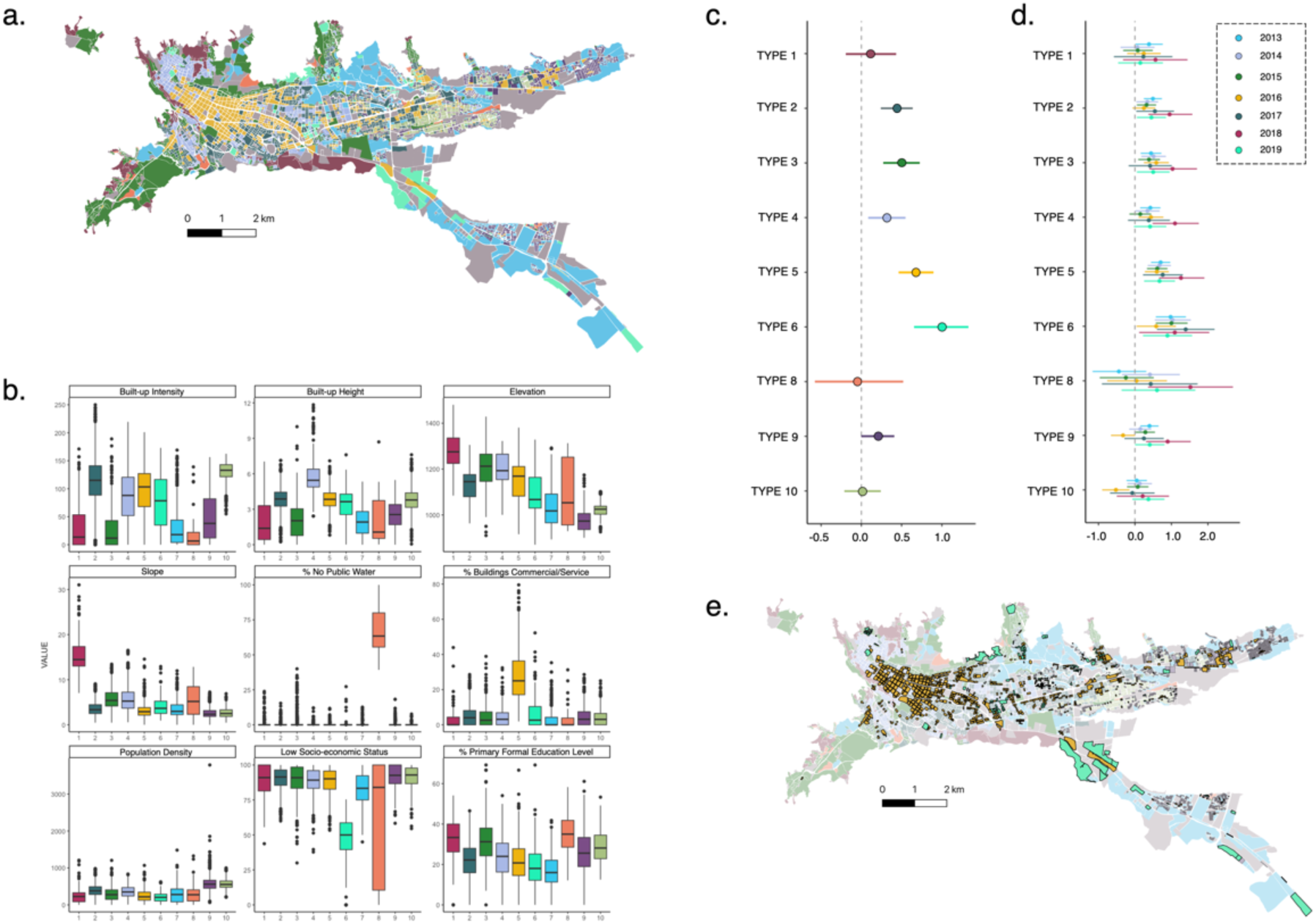
Associations between dengue incidence and landscape composition and configuration (census block scale) **a.** The 10 socio-environmental typologies mapped onto the census blocks of Ibagué. Type 1 census blocks are characterized as low-density blocks located in the urban periphery. Type 2 blocks are characterized as having high-population density (located in the urban core). Type 3 blocks are characterized as intermediate-density blocks (located in the urban periphery). Type 4 blocks are high-elevation high population/building density areas (located in the urban core). Type 5 are high-density commercial zones (located in the urban core). Type 6 are high socio-economic status and commercial areas (mixed urban core and periphery). Type 7 urban sections are intermediate socio-economic status (mixed urban core and periphery). Type 8 are low-serviced residential areas located in the urban periphery (low levels of formal water and education access). Type 9 census blocks are high-density areas located in the urban periphery. Type 10 are high-density residential areas located in the urban periphery. Full characterization of the 10 census block typologies are located in Table 3.2. **b.** Boxplots comparing the statistical values for the nine socio-environmental covariates retained in the hierarchical clustering algorithm. **c.** Coefficient plot showing the effect of nine typologies on total dengue cases during January 2013–July 2019 with 95% confidence intervals, compared to Typology 7 as a reference variable and offset by the population size of the census block. **d.** Coefficient plot showing the effect of nine typologies on annual dengue cases during 2013–2019 with 95% confidence intervals, compared to Typology 7 as a reference variable and offset by the population size of the census block. **e.** Map of Typologies 5 and 6, which have the highest positive statistical association with total dengue incidence, compared to reference Typology 7.

**Table 1.** Characterization of the 10-class typology at the urban section level.

| Typology | 1 | 2 | 3 | 4 | 5 | 6 | 7 | 8 | 9 | 10 |
| --- | --- | --- | --- | --- | --- | --- | --- | --- | --- | --- |
| Elevation | High | High | High | Int | Int | Int | Int | Low | Low | Mixed |
| Slope | High | Int | Int | Low | Low | Low | Low | Low | Low | Mixed |
| Built-Up Intens. | Mixed | Mixed | Int | High | High | Int | High | Low | High | Low |
| Commercial | Low | Low | Low | Low | Int | Low | Low | Low | High | Low |
| Pop. Density | Low | Int | Int | High | High | Int | High | Low | Int | Int |
| % Low-Estrato | High | High | Low | High | Low | Mixed | High | Low | Low | High |
| % Water Access | Int | Low | Low | Low | Low | Low | Low | Low | Low | High |
| Unocc. Housing | Int | Int | High | Int | Int | Int | Int | High | Int | Low |
| Core vs. Periphery | Periphery | Mixed | Mixed | Core | Core | Mixed | Core | Periphery | Core | Periphery |
|  | *High-elevation*<br>*Periphery* | *High-elevation*<br>*Low SES* | *High-elevation*<br>*High SES* | *High-elevation*<br>*High Density* | *High Density*<br>*Mixed-Use* | *Low Density* | *High Density*<br>*Residential* | *Low Density*<br>*Periphery* | *El Centro Commercial*<br>*District* | *Low-SES/*<br>*Periphery* |

**Table 2.** Characterization of the 10-class typology at the census block level.

| Typology | 1 | 2 | 3 | 4 | 5 | 6 | 7 | 8 | 9 | 10 |
| --- | --- | --- | --- | --- | --- | --- | --- | --- | --- | --- |
| Elevation | High | Int | High | High | Int | Mixed | Mixed | Mixed | Low | Low |
| Slope | High | Int | Int | Int | Low | Mixed | Mixed | Mixed | Low | Low |
| Built-Up Intens. | Low | High | Low | High | High | High | Int | Low | Int | High |
| Built-Up Height | Low | High | Low | High | High | High | Int | Low | Int | High |
| Commercial | Low | Int | Int | Int | High | High | Low | Low | Int | Int |
| Pop. Density | Low | High | Int | High | Int | Low | Int | Low | High | High |
| % Low-Estrato | High | High | High | High | High | Low | Int | Mixed | High | High |
| % Water Access | Mixed | High | Mixed | High | High | High | High | Low | High | High |
| % Formal Edu. | Int | Int | Int | Int | Int | Int | High | Low | Int | Int |
| Core vs. Periphery | Periphery | Core | Periphery | Core | Core | Mixed | Mixed | Periphery | Periphery | Core |
|  | *Low-density*<br>*periphery* | *High-density:*<br>*population*<br>*density* | *Int.-density*<br>*periphery* | *High-elevation*<br>*&*<br>*high-density*<br>*development* | *Core*<br>*commercial*<br>*zone* | *High SES &*<br>*commercial* | *Int. SES &*<br>*High*<br>*Formal*<br>*Education* | *Low-service*<br>*residential:*<br>*periphery* | *High-density*<br>*periphery* | *High-density*<br>*residential* |

**Table 3.** Negative binomial regression of urban section typologies.

| Factor | Estimate | SE | p-value | IRR (95% CI) |
| --- | --- | --- | --- | --- |
| **Intercept** | -4.217 | 0.421 | p<0.01 | 0.01 (0.01, 0.04) |
| Typology 8 | -- | -- | -- | -- |
| Typology 1 | 0.935 | 0.581 | 0.11 | 2.55 (0.81, 8.01) |
| Typology 2 | 1.125 | 0.442 | p<0.01* | 3.50 (1.36, 7.61) |
| Typology 3 | 0.963 | 0.451 | 0.03* | 2.62 (1.01, 5.80) |
| Typology 4 | 1.242 | 0.460 | p<0.01* | 3.46 (1.31, 7.86) |
| Typology 5 | 0.974 | 0.443 | 0.03* | 2.65 (1.03, 5.74) |
| Typology 6 | 0.860 | 0.581 | 0.05 | 2.36 (0.92, 5.17) |
| Typology 7 | 0.907 | 0.434 | 0.04* | 2.48 (0.98, 5.28) |
| Typology 9 | 1.468 | 0.477 | p<0.01* | 4.34 (1.61, 10.28) |
| Typology 10 | 0.715 | 0.613 | 0.24 | 2.04 (0.62, 7.07) |

**Table 4.** Negative binomial regression of urban section typologies.

| Factor | Estimate | SE | p-value | IRR (95% CI) |
| --- | --- | --- | --- | --- |
| **Intercept** | -3.58 | 0.082 | p<0.01 | 0.03 (0.01, 0.04) |
| Typology 7 | -- | -- | -- | -- |
| Typology 1 | 0.116 | 0.158 | 0.46 | 1.23 (0.81, 8.01) |
| Typology 2 | 0.442 | 0.099 | p<0.01* | 1.56 (1.36, 7.61) |
| Typology 3 | 0.500 | 0.113 | p<0.01* | 1.65 (1.31, 2.06) |
| Typology 4 | 0.318 | 0.114 | p<0.01* | 1.37 (1.10, 1.72) |
| Typology 5 | 0.678 | 0.107 | p<0.01* | 1.97 (1.59, 2.43) |
| Typology 6 | 0.984 | 0.168 | p<0.01* | 2.68 (1.93, 3.75) |
| Typology 8 | -0.057 | 0.280 | 0.84 | 0.94 (0.56, 1.67) |
| Typology 9 | 0.201 | 0.101 | 0.04 | 1.22 (1.00, 1.49) |
| Typology 10 | 0.013 | 0.112 | 0.91 | 1.01 (0.81, 1.27) |

#### Landscape configuration metrics

The best-fit multivariate model of total dengue incidence showed negative associations with Shannon’s diversity index within a 500-meter buffer (IRR = 0.58, p < 0.01), and negative associations with interspersion at the same distance. Increases in perimeter-area fractal dimension, indicating shape complexity, were positively associated with dengue incidence within a 1-km buffer (IRR = 1.37, p < 0.01).

## 4. Discussion

Dengue risk varies across cities due to heterogeneities in biophysical, socio-economic, institutional, and cultural properties (Kache et al., 2022). This study demonstrated that emergent patterns of dengue risk can be mapped using multidimensional characterizations of urban landscape composition, considering the spatial configuration of the multidimensional typologies. For the tropical Andean city of Ibagué, Colombia, dengue incidence was associated with typologies of landscape composition captured by biophysical heterogeneity (elevation, slope, built-up intensity), socio-economic variation (commercial building use, low socio-economic status housing, population density), and institutional heterogeneity (access to public water systems). Additionally, the diversity and aggregation versus patchiness of typologies played a significant role in understanding how local spatial relationships drive dengue risk.

By analyzing two geographic levels—the urban section and the census block—we pinpointed variables crucial for defining landscape composition at both scales, such as elevation and slope. Additionally, some variables only added value in the multi-dimensional parameter space for one specific scale. For instance, when assessing development levels within the city, principal component analysis showed that at the urban section level, "built-up intensity” effectively represented infrastructural variation citywide. In contrast, at the census block level, "built-up height" provided enough variation to be treated as an independent feature in hierarchical clustering.

Our multi-scale approach effectively identified areas where dengue risk was either spatially consistent or inconsistent across different scales. For instance, analysis at the urban section level showed that the typology associated with “El Centro” had dengue incidence rates 1.3 times those of the reference typology group at both the urban section and census block levels. El Centro has relatively high socioeconomic status housing and moderate residential population density; therefore, it is not considered a high-risk area for dengue in the city. However, as the main commercial district of Ibagué, it functions as a major daytime activity hub and a center for population mixing. These findings parallel the results of a socio-economic typology analysis conducted by Telle et al. in New Delhi, India, where centrally-located “rich units” exhibited higher dengue incidence rates compared to low-density, low socio-economic status units within the city, as well as previous research emphasizing the significance of commercial zones in relation to city-level dengue risk (Cavalcante et al., 2013; Dom et al., 2016; Telle et al., 2016b). In our detailed analysis using census blocks, Type 5 blocks associated with El Centro demonstrated the second-highest risk of dengue, following Type 6 blocks. Unlike Type 5, Type 6 blocks were spatially disaggregated, predominantly situated in the eastern half of the city. These blocks represented areas characterized by high socio-economic status and moderate population density, despite exhibiting intermediate to high levels of built-up intensity and building heights. Notably, buildings within these blocks exhibited high levels of non-residential usage, ranking second only to Type 5 blocks. An examination of sample pixel sites in Google Earth imagery and census data confirmed that Type 6 blocks were commercial zones, likely serving as hubs of local, neighborhood-level activity.

The observation that commercial spaces have the highest levels of dengue incidence in Ibagué is significant in several respects. First, this finding illustrates the importance of accounting for joint effects and interactions of multiple determinants of dengue risk in statistical approaches. For instance, as a single variable, the proportion of buildings with non-residential commercial use were not able to explain observed patterns of dengue incidence on its own, however when incorporated into a multi-dimensional characterization of the urban landscape, appear to drive? a significant positive effect on dengue risk. Second, the high levels of dengue incidence observed within commercial zones indicate that vector monitoring and control activities should be systematically expanded beyond residential areas into high-activity areas and blocks of the city, as was initiated by the Secretary of Health of Ibagué on a small scale in 2019 but subsequently discontinued (Ho et al., 2005). Third, *El Centro* represents the oldest Spanish colonial neighborhood within Ibagué, constructed during the sixteenth century (Francel, 2017). Therefore, while building age was not available for consideration in this analysis, the deterioration of the built infrastructure due to its building age, may contribute to higher densities of *Aedes* spp. larval habitat within the neighborhood or higher levels of mosquito-human exposure, and require further field-based exploration (Sun et al., 2021). Finally, these results indicate that further research is warranted, examining mobility fluxes between peripheral, residential neighborhoods and *El Centro*. Population cell phone data, for example, may provide insight into the spatio-temporal dynamics of dengue introduction within Ibagué (Stoddard et al., 2009; Vazquez-Prokopec et al., 2013). Additionally, mechanistic meta-population models parameterized with real-world data, can advance theoretical understandings of the relative role of landscape composition, configuration and connectivity in driving dengue risk and transmission (Colizza and Vespignani, 2008; Kache et al., 2022; Lee and Castillo-Chavez, 2015b).

Alongside landscape composition, we analyze how configuration influences risk. High diversity in typologies and low patch aggregation in surrounding buffer areas of a given spatial unit protect against dengue incidence at both the census block and urban section levels. These findings imply that having socio-environmentally diverse census blocks and urban sections nearby offers city-wide benefits, while large contiguous patches of uniform typology pose disadvantages. This aligns with existing U.S. research on neighborhood effects on well-being, where “social mixing” and the presence of wealthier neighbors seem to create positive externalities for less affluent neighbors, possibly through social leverage and influence involving higher-level social actors within the urban system (Kache et al., 2022; Van Ham et al., 2011). In

Australia, studies show that large neighborhoods with uniform housing and socio-economically disadvantaged tenants reinforce socio-spatial exclusion and neighborhood stigmatization. Conversely, in Argentina, neighborhoods with highly mixed socio-economic compositions tend to have less social cohesion, which is important for community-based dengue prevention efforts (Sánchez et al., 2007; Stewart Ibarra et al., 2014). Likely, both mechanisms operate at different scales and areas within Ibagué but need further confirmation through household and neighborhood socio-ecological surveys (Arthurson, 2010; Sánchez et al., 2007). Additionally, mixed-methods geographic approaches should be employed to understand the spatial dynamics of “social mixing” and dengue risk, distinguishing between social mixing within neighborhoods as colloquially understood versus proximity-based social mixing relating to our study.

Patch shape also had a scale-dependent relationship with dengue risk. Increases in shape complexity at the census block scale were positively associated with dengue incidence; however, this association was not significant at the urban section scale. We hypothesize that shape complexity at a census block scale captures areas of the city that were informally developed without local regulatory oversight, reflecting land parcels of inconsistent shape and size. These findings are consistent with geospatial analyses conducted at the building scale by Sarfraz et al., who found that dengue cases tended to cluster around irregularly-shaped structures located near roadways (Sarfraz et al., 2014). However, shape-based metrics have been an under-explored tool in geospatial studies of dengue risk, although they have been separately implemented in studies of malaria risk (Marti et al., 2020). Uncovering the mechanisms by which landscape configuration affects *Aedes* spp. hazard and exposure is a necessary precursor to understanding how urban expansion will alter the spatial epidemiology of *Aedes*-borne diseases; and requires characterization of the landscape that go beyond the dichotomy of urban versus non-urban land cover classes (Kache et al., 2022; Little et al., 2011b; Troyo et al., 2009).

The geographic units of analysis leveraged in this study are established at the national level in Colombia. They were selected based on their compatibility with national census data and to minimize loss of information when aggregating socio-economic data to multiple geographic boundaries. The degree to which these administrative boundaries correspond to official and unofficial neighborhood boundaries within Ibagué is beyond the scope of this analysis. In the broader effort to characterize the “building blocks” of urban landscapes, census blocks and urban sections may not fully reflect bottom-up local processes within Ibagué. Therefore, future field-based efforts should be used to ground-truth these typologies and generate information regarding the cultural and institutional properties of the urban system to redefine typology assignments. Nevertheless, our approach operationalizes a hierarchical patch systems framework for dengue risk proposed by Kache et al. (2022) using remotely-sensed and open access census data, encouraging the replicability of this approach for cities worldwide.

## Data Availability

All data produced in the present study are available upon reasonable request to the authors

## References

Alberti, M., Wang, T., 2022. Detecting patterns of vertebrate biodiversity across the multidimensional urban landscape. Ecology Letters 25, 1027–1045.

Alto, B.W., Juliano, S.A., 2001. Temperature effects on the dynamics of Aedes albopictus (Diptera: Culicidae) populations in the laboratory. Journal of medical entomology 38, 548–556.

Aronson, M.F., Lepczyk, C.A., Evans, K.L., Goddard, M.A., Lerman, S.B., MacIvor, J.S., Nilon, C.H., Vargo, T., 2017. Biodiversity in the city: key challenges for urban green space management. Frontiers in Ecology and the Environment 15, 189–196.

Arthurson, K., 2010. Operationalising social mix: Spatial scale, lifestyle and stigma as mediating points in resident interaction. Urban policy and research 28, 49–63.

Carrasquilla, M.C., Ortiz, M.I., León, C., Rondón, S., Kulkarni, M.A., Talbot, B., Sander, B., Vásquez, H., Cordovez, J.M., González, C., 2021. Entomological characterization of Aedes mosquitoes and arbovirus detection in Ibagué, a Colombian city with co-circulation of Zika, dengue and chikungunya viruses. Parasites & vectors 14, 1–14.

Cavalcante, M.P.R., Oliveira, C. de, Simão, F.B., Lima, P.R., Monteiro, P.S., 2013. Geospatial analysis: a study about dengue. Acta Paulista de Enfermagem 26, 360–368.

Colizza, V., Vespignani, A., 2008. Epidemic modeling in metapopulation systems with heterogeneous coupling pattern: Theory and simulations. Journal of theoretical biology 251, 450–467.

DANE, 2018. Censo Nacional de Población y Vivienda, 2018.

Deng, J.S., Wang, K., Hong, Y., Qi, J.G., 2009. Spatio-temporal dynamics and evolution of land use change and landscape pattern in response to rapid urbanization. Landscape and urban planning 92, 187–198.

Desa, U., 2018. Revision of world urbanization prospects. UN Department of Economic and Social Affairs 16.

Dimoudi, A., Kantzioura, A., Zoras, S., Pallas, C., Kosmopoulos, P., 2013. Investigation of urban microclimate parameters in an urban center. Energy and Buildings 64, 1–9.

Dom, N.C., Ahmad, A.H., Abd Latif, Z., Ismail, R., 2016. Application of geographical information system-based analytical hierarchy process as a tool for dengue risk assessment. Asian Pacific Journal of Tropical Disease 6, 928–935.

Dunn†, J.C., 1974. Well-Separated Clusters and Optimal Fuzzy Partitions. Journal of Cybernetics 4, 95–104. 10.1080/01969727408546059

Evans, M.V., Hintz, C.W., Jones, L., Shiau, J., Solano, N., Drake, J.M., Murdock, C.C., 2019. Microclimate and larval habitat density predict adult Aedes albopictus abundance in urban areas. The American journal of tropical medicine and hygiene 101, 362.

Fikrig, K., Peck, S., Deckerman, P., Dang, S., St Fleur, K., Goldsmith, H., Qu, S., Rosenthal, H., Harrington, L.C., 2020. Sugar feeding patterns of New York Aedes albopictus mosquitoes are affected by saturation deficit, flowers, and host seeking. PLoS Neglected Tropical Diseases 14, e0008244.

Foot, D., 2017. Operational urban models: an introduction. Routledge.

Fragkias, M., Güneralp, B., Seto, K.C., Goodness, J., 2013. A synthesis of global urbanization projections. Urbanization, biodiversity and ecosystem services: Challenges and opportunities 409–435.

Francel, A., 2017. La superposición de cartografía histórica como método de análisis morfológico y toma de decisiones urbanísticas. Ibagué, Colombia, 1935-2016. urbe. Revista Brasileira de Gestão Urbana 9, 293–313.

García-Betancourt, T., Higuera-Mendieta, D.R., González-Uribe, C., Cortés, S., Quintero, J., 2015. Understanding water storage practices of urban residents of an endemic dengue area in Colombia: Perceptions, rationale and socio-demographic characteristics. PloS one 10, e0129054.

Grove, J.M., Locke, D.H., O’Neil-Dunne, J.P., 2014. An ecology of prestige in New York City: Examining the relationships among population density, socio-economic status, group identity, and residential canopy cover. Environmental management 54, 402–419.

Harrington, L.C., Scott, T.W., Lerdthusnee, K., Coleman, R.C., Costero, A., Clark, G.G., Jones, J.J., Kitthawee, S., Kittayapong, P., Sithiprasasna, R., 2005. Dispersal of the dengue vector Aedes aegypti within and between rural communities. The American journal of tropical medicine and hygiene 72, 209–220.

Hemme, R.R., Thomas, C.L., Chadee, D.D., Severson, D.W., 2010. Influence of urban landscapes on population dynamics in a short-distance migrant mosquito: evidence for the dengue vector Aedes aegypti. PLoS Neglected Tropical Diseases 4, e634.

Ho, C.-M., Feng, C.-C., Yang, C.-T., Lin, M.-W., Teng, H.-C., Mhl, L.T., Hsu, E., Wu, S., Pai, H., Yin, C., 2005. Surveillance for dengue fever vectors using ovitraps at Kaohsiung and Tainan in Taiwan. Formosan Entomol 25, 159–174.

Honório, N.A., Silva, W. da C., Leite, P.J., Gonçalves, J.M., Lounibos, L.P., Lourenço-de-Oliveira, R., 2003. Dispersal of Aedes aegypti and Aedes albopictus (Diptera: Culicidae) in an urban endemic dengue area in the State of Rio de Janeiro, Brazil. Memórias do Instituto Oswaldo Cruz 98, 191–198.

Ibarra, A.M.S., Luzadis, V.A., Cordova, M.J.B., Silva, M., Ordoñez, T., Ayala, E.B., Ryan, S.J., 2014. A social-ecological analysis of community perceptions of dengue fever and Aedes aegypti in Machala, Ecuador. BMC public health 14, 1–12.

Kache, P.A., Santos-Vega, M., Stewart-Ibarra, A.M., Cook, E.M., Seto, K.C., Diuk-Wasser, M.A., 2022. Bridging landscape ecology and urban science to respond to the rising threat of mosquito-borne diseases. Nature Ecology & Evolution 1–16.

Kenneson, A., Beltrán-Ayala, E., Borbor-Cordova, M.J., Polhemus, M.E., Ryan, S.J., Endy, T.P., Stewart-Ibarra, A.M., 2017. Social-ecological factors and preventive actions decrease the risk of dengue infection at the household-level: Results from a prospective dengue surveillance study in Machala, Ecuador. PLoS neglected tropical diseases 11, e0006150.

Kropf, K., 2009. Aspects of urban form. Urban morphology 13, 105.

LaDeau, S.L., Allan, B.F., Leisnham, P.T., Levy, M.Z., 2015. The ecological foundations of transmission potential and vector-borne disease in urban landscapes. Functional ecology 29, 889–901.

Ledogar, R.J., Arosteguí, J., Hernández-Alvarez, C., Morales-Perez, A., Nava-Aguilera, E., Legorreta-Soberanis, J., Suazo-Laguna, H., Belli, A., Laucirica, J., Coloma, J., 2017. Mobilising communities for Aedes aegypti control: the SEPA approach. BMC Public Health 17, 103–114.

Lee, S., Castillo-Chavez, C., 2015. The role of residence times in two-patch dengue transmission dynamics and optimal strategies. Journal of theoretical biology 374, 152–164.

Leong, M., Dunn, R.R., Trautwein, M.D., 2018. Biodiversity and socioeconomics in the city: a review of the luxury effect. Biology Letters 14, 20180082.

Little, E., Barrera, R., Seto, K.C., Diuk-Wasser, M., 2011. Co-occurrence patterns of the dengue vector Aedes aegypti and Aedes mediovitattus, a dengue competent mosquito in Puerto Rico. Ecohealth 8, 365–375.

Marti, R., Li, Z., Catry, T., Roux, E., Mangeas, M., Handschumacher, P., Gaudart, J., Tran, A., Demagistri, L., Faure, J.-F., 2020. A mapping review on urban landscape factors of dengue retrieved from earth observation data, GIS techniques, and survey questionnaires. Remote Sensing 12, 932.

Michalos, A.C., 2014. Encyclopedia of quality of life and well-being research. Springer Netherlands Dordrecht.

Middel, A., Häb, K., Brazel, A.J., Martin, C.A., Guhathakurta, S., 2014. Impact of urban form and design on mid-afternoon microclimate in Phoenix Local Climate Zones. Landscape and urban planning 122, 16–28.

Mitchell-Foster, K.L., 2013. Interdisciplinary knowledge translation and evaluation strategies for participatory dengue prevention in Machala, Ecuador.

Mordecai, E.A., Cohen, J.M., Evans, M.V., Gudapati, P., Johnson, L.R., Lippi, C.A., Miazgowicz, K., Murdock, C.C., Rohr, J.R., Ryan, S.J., 2017. Detecting the impact of temperature on transmission of Zika, dengue, and chikungunya using mechanistic models. PLoS neglected tropical diseases 11, e0005568.

Murdock, C.C., Evans, M.V., McClanahan, T.D., Miazgowicz, K.L., Tesla, B., 2017. Fine-scale variation in microclimate across an urban landscape shapes variation in mosquito population dynamics and the potential of Aedes albopictus to transmit arboviral disease. PLoS neglected tropical diseases 11, e0005640.

Murtagh, F., Legendre, P., 2014. Ward’s hierarchical agglomerative clustering method: which algorithms implement Ward’s criterion? Journal of classification 31, 274–295.

Núñez Pita, J.M., 2021. Análisis Multitemporal de los cambios de la superficie de cultivo de arroz de la vereda Picaleña (Ibagué Tolima) sector rural entre los periodos 2010/2015/2020.

Pandey, B., Brelsford, C., Seto, K.C., 2022. Infrastructure inequality is a characteristic of urbanization. Proceedings of the National Academy of Sciences 119, e2119890119.

Plummer, R., de Loë, R., Armitage, D., 2012. A systematic review of water vulnerability assessment tools. Water resources management 26, 4327–4346.

Reiner Jr, R.C., Stoddard, S.T., Scott, T.W., 2014. Socially structured human movement shapes dengue transmission despite the diffusive effect of mosquito dispersal. Epidemics 6, 30–36.

Romeo-Aznar, V., Freitas, L.P., Cruz, O.G., King, A., Pascual, M., 2021. Fine-scale heterogeneity in population density predicts wave dynamics in dengue epidemics. medRxiv.

Rowley, W.A., Graham, C.L., 1968. The effect of temperature and relative humidity on the flight performance of female Aedes aegypti. Journal of Insect Physiology 14, 1251–1257.4

Roy Chowdhury, R., Larson, K., Grove, M., Polsky, C., Cook, E., Onsted, J., Ogden, L., 2011. A multi-scalar approach to theorizing socio-ecological dynamics of urban residential landscapes. Cities and the Environment (CATE) 4, 6.

Samson, D.M., Qualls, W.A., Roque, D., Naranjo, D.P., Alimi, T., Arheart, K.L., Müller, G.C., Beier, J.C., Xue, R.-D., 2013. Resting and energy reserves of Aedes albopictus collected in common landscaping vegetation in St. Augustine, Florida. Journal of the American Mosquito Control Association 29, 231.

Sánchez, D., Sassone, S., Matossian, B., 2007. Barrios y áreas sociales de San Carlos de Bariloche: análisis geográfico de una ciudad fragmentada. Presented at the IX Jornadas Argentinas de Estudios de Población, Asociación de Estudios de Población de la Argentina.

Sarfraz, M.S., Tripathi, N.K., Kitamoto, A., 2014. Near real-time characterisation of urban environments: a holistic approach for monitoring dengue fever risk areas. International Journal of Digital Earth 7, 916–934.

Scott, T.W., Morrison, A.C., 2010. Vector dynamics and transmission of dengue virus: implications for dengue surveillance and prevention strategies. Dengue virus 115–128.

Seto, K.C., Dhakal, S., Bigio, A., Blanco, H., Delgado, G.C., Dewar, D., Huang, L., Inaba, A., Kansal, A., Lwasa, S., 2014. Human settlements, infrastructure and spatial planning.

Seto, K.C., Sánchez-Rodríguez, R., Fragkias, M., 2010. The new geography of contemporary urbanization and the environment. Annual review of environment and resources 35, 167–194.

Stewart Ibarra, A.M., Luzadis, V.A., Borbor Cordova, M.J., Silva, M., Ordoñez, T., Beltran Ayala, E., Ryan, S.J., 2014. A social-ecological analysis of community perceptions of dengue fever and Aedes aegypti in Machala, Ecuador. BMC public health 14, 1–12.

Stoddard, S.T., Morrison, A.C., Vazquez-Prokopec, G.M., Paz Soldan, V., Kochel, T.J., Kitron, U., Elder, J.P., Scott, T.W., 2009. The role of human movement in the transmission of vector-borne pathogens. PLoS neglected tropical diseases 3, e481. 10.1371/journal.pntd.0000481

Stokes, E.C., Seto, K.C., 2019. Characterizing and measuring urban landscapes for sustainability. Environmental Research Letters 14, 045002.

Streutker, D.R., 2002. A remote sensing study of the urban heat island of Houston, Texas. International Journal of Remote Sensing 23, 2595–2608.

Sun, H., Dickens, B.L., Richards, D., Ong, J., Rajarethinam, J., Hassim, M.E., Lim, J.T., Carrasco, L.R., Aik, J., Yap, G., 2021. Spatio-temporal analysis of the main dengue vector populations in Singapore. Parasites & vectors 14, 1–11.

Telle, O., Vaguet, A., Yadav, N., Lefebvre, B., Daudé, E., Paul, R.E., Cebeillac, A., Nagpal, B., 2016. The spread of dengue in an endemic urban milieu–the case of Delhi, India. PloS one 11, e0146539.

Thapa, R.B., Murayama, Y., 2009. Examining spatiotemporal urbanization patterns in Kathmandu Valley, Nepal: Remote sensing and spatial metrics approaches. Remote Sensing 1, 534–556.

Troyo, A., Fuller, D.O., Calderón-Arguedas, O., Solano, M.E., Beier, J.C., 2009. Urban structure and dengue incidence in Puntarenas, Costa Rica. Singapore journal of tropical geography 30, 265–282.

Van Ham, M., Manley, D., Bailey, N., Simpson, L., Maclennan, D., 2011. Neighbourhood effects research: New perspectives, in: Neighbourhood Effects Research: New Perspectives. Springer, pp. 1–21.

Vazquez-Prokopec, G.M., Bisanzio, D., Stoddard, S.T., Paz-Soldan, V., Morrison, A.C., Elder, J.P., Ramirez-Paredes, J., Halsey, E.S., Kochel, T.J., Scott, T.W., 2013. Using GPS technology to quantify human mobility, dynamic contacts and infectious disease dynamics in a resource-poor urban environment. PloS one 8, e58802.

Whiteford, L.M., 1997. The ethnoecology of dengue fever. Medical anthropology quarterly 11, 202–223.

Wong, P.P.-Y., Lai, P.-C., Low, C.-T., Chen, S., Hart, M., 2016. The impact of environmental and human factors on urban heat and microclimate variability. Building and Environment 95, 199–208.

