## Supplementary material for "Socio-environmental typologies of dengue risk at multiple spatial scales of the urban landscape": SI

### Supplemental Methods

#### Data sources

***Dengue cases***

Using the national case definition, probable cases are classified based on either: 1) clinical diagnosis with two or more symptoms and at least one serological immunoglobulin M (IgM) test; or 2) an epidemiological link to a confirmed case within 14 days of symptom onset. Confirmed dengue cases are identified through laboratory diagnosis via 1) PCR within five days of symptom onset; or 2) IgM serology using Dengue ELISA kits after five days of symptom onset. During an outbreak, serological samples are collected from 5% of dengue fever cases and from all severe dengue cases.

***Socio-environmental data and spatial layers***

We analyzed data for 4,999 residential census blocks (i.e., at least one dwelling reported in the 2018 census) out of 5,571 total census blocks in the city. Residential census blocks (hereafter, census blocks) had a median spatial extent of .2 hectares per block [Interquartile range (IQR): 0.12–0.43 hectares]. Additionally, we examined 269 residential urban sections out of 271 total sections, with a median extent of 10.2 hectares per section (IQR: 6.6–18.3 hectares).

#### Typology analysis

The resulting tree is visualized as a dendrogram. We used the average silhouette approach to measure how well each observation lies within each cluster [107]. This method calculates the average silhouette for each observation at different values of *k* clusters. The optimal number of clusters maximizes the average silhouette value over a range of possible *k* clusters.

The stability measures evaluate the stability of a clustering result by comparing it with the clusters obtained by removing one column at a time. These measures include the average proportion of non-overlap (APN), the average distance (AD), the average distance between means (ADM), and the figure of merit (FOM). The APN, AD, and ADM are all based on the cross-classification table of the original clustering with the clustering based on the removal of one column. The APN measures the average proportion of observations not placed in the same cluster under both cases, while the AD measures the average distance between observations placed in the same cluster under both cases and the ADM measures the average distance between cluster centers for observations placed in the same cluster under both cases. The FOM measures the average intra-cluster variance of the deleted column, where the clustering is based on the remaining (undeleted) columns. In all cases the average is taken over all the deleted columns, and all measures should

### Tables

**Table S.1 Socio-environmental candidate variables for consideration in typology analysis**

| Urban Property | Variable | Description & Justification | Data Source & Processing |
|---|---|---|---|
| Biophysical | Normalized Difference Vegetation Index (NDVI) | Calculated by measuring difference between near-infrared band (which vegetation strongly reflects) and red band (which vegetation absorbs). Values range from -1 (less vegetation) to +1 (more vegetation). | **Source:** Landsat 8 (30-meters)<br>**Functions/Tools:** Cloud-free composite [Google Earth Engine (GEE)], NDVI; Zonal Statistics, Median Pixel Value (ArcGIS) |
| | Elevation | Median elevation across all pixels within the neighborhood. Derived from SRTM digital elevation model (DEM). | **Source:** Shuttle Radar Topography Mission (SRTM) (30-meters), NASA Jet Propulsion Laboratories.<br>**Functions/Tools:** Zonal Statistics, Median Pixel Value (ArcGIS) |
| | Slope | Measures the rate of change of elevation for each DEM cell, calculated in degrees. The values range from 0 to 90. | **Source:** SRTM (30-meters), NASA Jet Propulsion Laboratories.<br>**Functions/Tools:** Zonal Statistics, Median Pixel Value (ArcGIS) |
| | Built-Up Intensity (BUI) | Measures built-up areas based on supervised classification (using spatial, textural data) of Landsat imagery. Values range from 0 (not built-up) to 255 (entirely built up). | **Source:** Global Human Settlements Layer (GHSL) (30-meters)<br>**Functions/Tools:** Zonal Statistics, Median Pixel Value (ArcGIS) |
| | Built-Up Height | Spatial distribution of the building heights as extracted from the filtering of a composite of global digital elevation models (DEM). | **Source:** Global Human Settlements (GHS-BUILT-H) (100-meters)<br>**Functions/Tools:** Zonal Statistics, Median Pixel Value (ArcGIS) |
| | Street Nodal Density | We divided the number of street intersection nodes by the area of the geographic unit. At the census block level, a 25-meter buffer was created around the polygon to calculate the density. | **Source:** Open Street Map<br>**Functions/ Tools:** Network Analyst, Buffer, Point-in-Polygon (ArcGIS) |
| Institutional | Proportion Water System Access | The proportion of dwellings with access to the public water system | |

| | | | |
|---|---|---|---|
| | Proportion Sewage System Access | The proportion of dwellings with access to the public sewage system | **Source:** Colombia National Census (Departamento Administrativo Nacional de Estadística, DANE), Microdata, 2018 |
| | Proportion Trash Collection Access | The proportion of dwellings with trash collection | |
| **Socio-Economic** | Proportion of Buildings with Service or Commercial Use | The proportion of buildings with a non-residential service or commercial use | |
| | Proportion of Units Unoccupied | The proportion of residential units that are not occupied with tenants | |
| | Population Density | The number of persons per $100m^2$ | |
| | Within-household Occupant Density | The average number of occupants per household | |
| | % Population Under 10 Years | The proportion of persons under 10 years of age. | |
| | % Internet | The percent of dwellings with internet access | |
| | % Highest Schooling Attained: Primary School | The percent of the population that is able to read/write in Spanish | |
| | % *Estratos* 1[§] | Proportion of residential buildings that have an *Estrato* 1 designation. | |

[§] Households across all Colombian cities are divided into socio-economic strata (i.e., *Estratos*) for tax and utilities purpose. For *Estratos* 1–3, citizens receive subsidies in services (water, gas, electricity). The highest *Estratos* 5–6, contribute to lower-*Estrato* subsidies with bills higher than their consumption. To designate the *Estrato* of a building, the municipality government evaluates the façade of its house, the building and roofing materials, and the conditions of the road in front of the house. The model does not incorporate income, number of people who make up a family, age, or if household members have any type of disability or employment.

**Table S.2 Landscape metrics used to calculate configuration variables**

| Category | Acronym | Metric name | Description |
|---|---|---|---|
| **Aggregation** | COHESION | Patch cohesion index | Connectedness of patches |
| | PLADJ | Percent of Like Adjacencies | Calculates the frequency how often patches of different classes *i* (focal class) and *k* are next to each other. Equals PLADJ = 0 if class *i* is maximal disaggregated, i.e. every cell is a different patch. Equals PLADJ = 100 when the only one patch is present. |
| | CONTAG | Effective mesh size | The probability of two random cells within the landscape belonging to the same class, based on cell adjacencies. Affected by the dispersion and interspersion of classes. E.g., low class dispersion (= high proportion of like adjacencies) and low interspersion (= uneven distribution of pairwise adjacencies) lead to a high contagion value. |
| **Diversity & Complexity** | ENT | Marginal entropy | Marginal entropy is the average total information provided by observing two variables "X", and "Y" |
| | SHDI | Shannon's Diversity Index | Calculates the diversity, taking the number of different classes and the abundance of each class into account. SHDI = 0 when only one patch is present and increases, without limit, as the number of classes increases while the proportions are equally distributed |
| | PRD | Patch Richness Density | Calculates the number of classes per total landscape area in $m^2$. Approaches PRD > 1 when one patch is present and the landscape is rather large. Increases, without limit, as the number of classes increases and the landscape area decreases. |

| | | | |
|---|---|---|---|
| **Shape** | PAFRAC | Perimeter-Area Fractal Dimension | Describes the patch complexity of the landscape while being scale independent. This means that increasing the patch size while not changing the patch form will not change the metric. |
| | FRAC_MN | Mean fractal dimension index | Summarizes the landscape as the mean of the fractal dimension index of all patches in the landscape. The fractal dimension index is based on the patch perimeter and the patch area and describes the patch complexity. |

**Table S.3 Principal components (PC) analysis loadings for the urban section and census block**

| Spatial Scale | Variable | PC1 | PC2 | PC3 | PC4 | PC5 |
|---|---|---|---|---|---|---|
| **Urban Section** | Elevation | 20.50 | 0.00 | 29.87 | 0.29 | 0.86 |
| | Slope | 17.93 | 13.43 | 8.02 | 15.41 | 3.78 |
| | Built-Up Intensity (BUI) | 8.71 | 13.82 | 22.02 | 0.15 | 14.87 |
| | % Water System Access | 1.35 | 13.40 | 0.11 | 62.06 | 21.96 |
| | % of Buildings with Service Use | 4.67 | 27.84 | 6.75 | 8.30 | 0.06 |
| | Proportion of Units Unoccupied | 6.31 | 4.94 | 25.89 | 10.19 | 44.91 |
| | Population Density | 31.65 | 0.10 | 2.40 | 1.05 | 12.67 |
| | % *Estratos* 1 | 8.88 | 26.47 | 4.93 | 2.54 | 0.89 |
| | Elevation | 27.33 | 13.25 | 5.01 | 0.19 | 1.05 |
| | Slope | 29.20 | 0.03 | 17.91 | 0.40 | 2.77 |

| | | | | | | |
|---|---|---|---|---|---|---|
| **Census Block** | Built-Up Intensity (BUI) | 7.52 | 22.08 | 2.69 | 13.68 | 0.13 |
| | Built-Up Height | 1.32 | 29.53 | 3.48 | 11.45 | 2.99 |
| | % Water System Access | 6.00 | 6.59 | 1.49 | 45.52 | 1.69 |
| | % of Buildings with Service Use | 0.33 | 15.53 | 11.82 | 0.09 | 46.02 |
| | Population Density | 23.53 | 3.78 | 11.65 | 2.36 | 2.34 |
| | % Primary-Level Formal Edu. | 0.78 | 8.41 | 19.36 | 10.18 | 27.96 |
| | % *Estratos* 1 | 3.99 | 0.79 | 26.58 | 16.24 | 15.06 |

**Table S.4 Internal and stability cluster validation measure for hierarchical clustering algorithm**

| Number of clusters (k) | **Urban Section** | | | | | **Census Block** | | | | |
|---|---|---|---|---|---|---|---|---|---|---|
| | Dunn Index | APN | AD | ADM | FOM | Dunn Index | APN | AD | ADM | FOM |
| **2** | 0.068 | **0.221**§ | 3.441 | 0.806 | 0.978 | 0.030 | **0.404**§ | 3.838 | **1.108**§ | 0.994 |
| **3** | 0.068 | 0.227 | 3.158 | **0.775**§ | 0.945 | 0.031 | 0.445 | 3.702 | 1.184 | 0.975 |
| **4** | 0.068 | 0.253 | 3.013 | 0.932 | 0.932 | 0.031 | 0.470 | 3.586 | 1.207 | 0.962 |
| **5** | 0.068 | 0.246 | 2.823 | 0.809 | 0.915 | 0.032 | 0.536 | 3.533 | 1.306 | 0.958 |
| **6** | 0.105 | 0.275 | 2.723 | 0.840 | 0.910 | **0.032**§ | 0.517 | 3.424 | 1.277 | 0.951 |
| **7** | **0.118**§ | 0.337 | 2.638 | 0.911 | 0.907 | 0.032 | 0.533 | 3.355 | 1.271 | 0.950 |
| **8** | 0.112 | 0.327 | 2.541 | 0.889 | 0.898 | 0.032 | 0.562 | 3.306 | 1.300 | 0.948 |
| **9** | 0.112 | 0.348 | 2.482 | 0.900 | 0.891 | 0.032 | 0.590 | 3.263 | 1.316 | 0.945 |
| **10** | 0.112 | 0.363 | **2.408**§ | 0.895 | **0.885**§ | 0.025 | 0.580 | **3.213**§ | 1.288 | **0.940***|

§ Indicates the optimal number of typologies based on internal validity and stability metrics

**Table S.5 Model Comparison For Urban Section-level Configuration Metrics**

| Metric Class | Landscape Metric | Buffer Distance | AIC |
|---|---|---|---|
| **Aggregation** | **CONTAG** | 0.5 km | 2743.6 |
| | | 1.0 km | 2752.0 |
| | | 2.0 km | 2743.1 |
| | **PLADJ** | 0.5 km | 2745.2 |
| | | 1.0 km | 2749.2 |
| | | 2.0 km | 2751.6 |

| | | | |
|---|---|---|---|
| **Diversity** | **ENT** | 0.5 km | 2750.6 |
| | | 1.0 km | 2745.1 |
| | | 2.0 km | 2751.6 |
| | **SHDI** | 0.5 km | 2750.7 |
| | | 1.0 km | 2745.1 |
| | | 2.0 km | 2741.1 |
| | **PRD** | 0.5 km | 2752.0 |
| | | 1.0 km | 2750.5 |
| | | 2.0 km | 2749.5 |
| **Shape** | **PAFRAC** | 0.5 km | 2741.9 |
| | | 1.0 km | 2752.1 |
| | | 2.0 km | 2751.7 |
| | **MEAN FRAC** | 0.5 km | 2749.1 |
| | | 1.0 km | 2747.2 |
| | | 2.0 km | 2743.7 |

### Figures

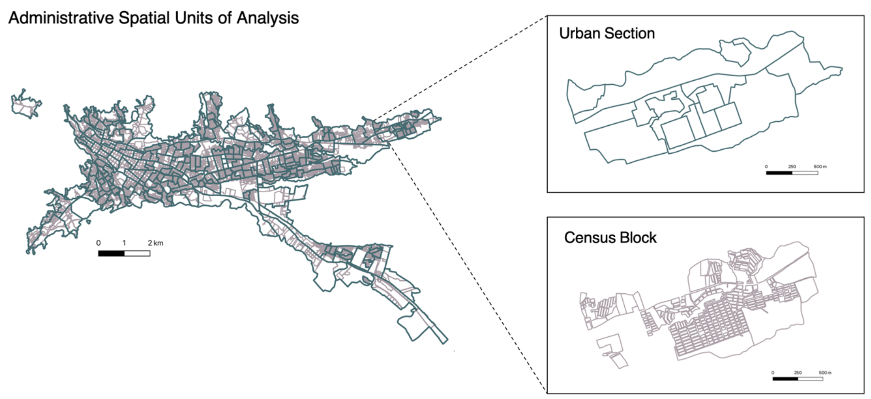


**Figure S.1 Map of administrative spatial units of analysis for the study**

We analyzed data for 4,999 residential census blocks (with at least one dwelling reported) out of the 5,571 total census blocks that make up the city. Residential census blocks had a median spatial extent of .2 hectares per block [Interquartile range (IQR): 0.12–0.43 hectares]. Additionally, we examined data for 269 out of 271 total urban sections, which have a median extent of 10.2 hectares per section (IQR: 6.6–18.3 hectares).

**Commented [MOU1]:** can you draw the outlines of urban sections on the 'census block' figure to see how they are nested?

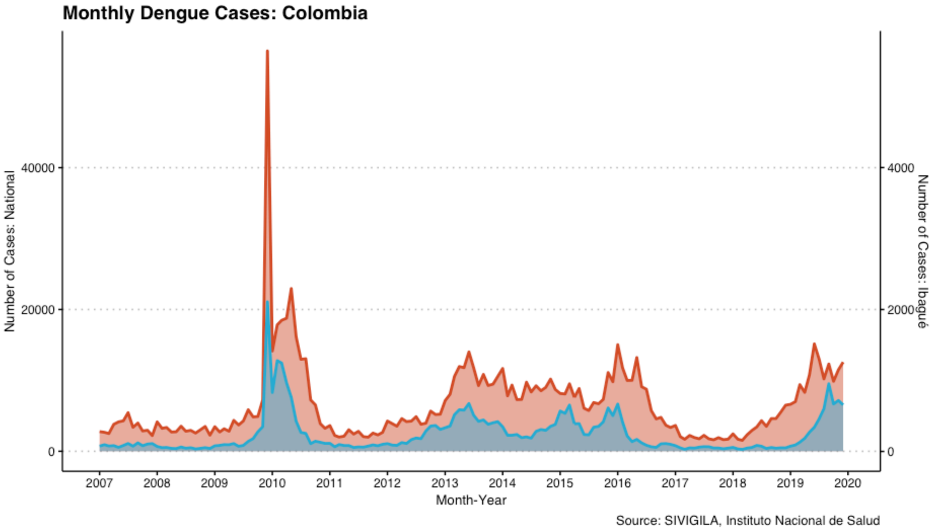


**Figure S.2 Monthly number of cases in Ibagué, Colombia compared to national case counts**

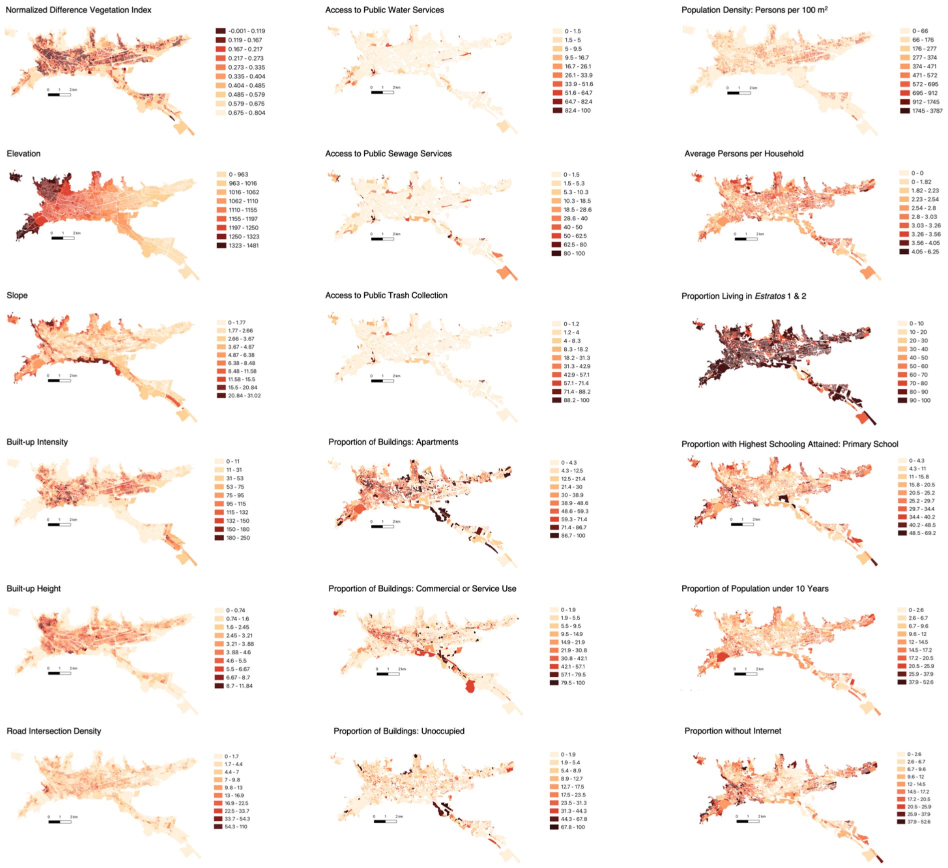


**Figure S.3 Maps of socio-environmental candidate variables at the urban section level**

Choropleth maps are shown with Jenks natural breaks to optimize the arrangement of values into "natural" classes.

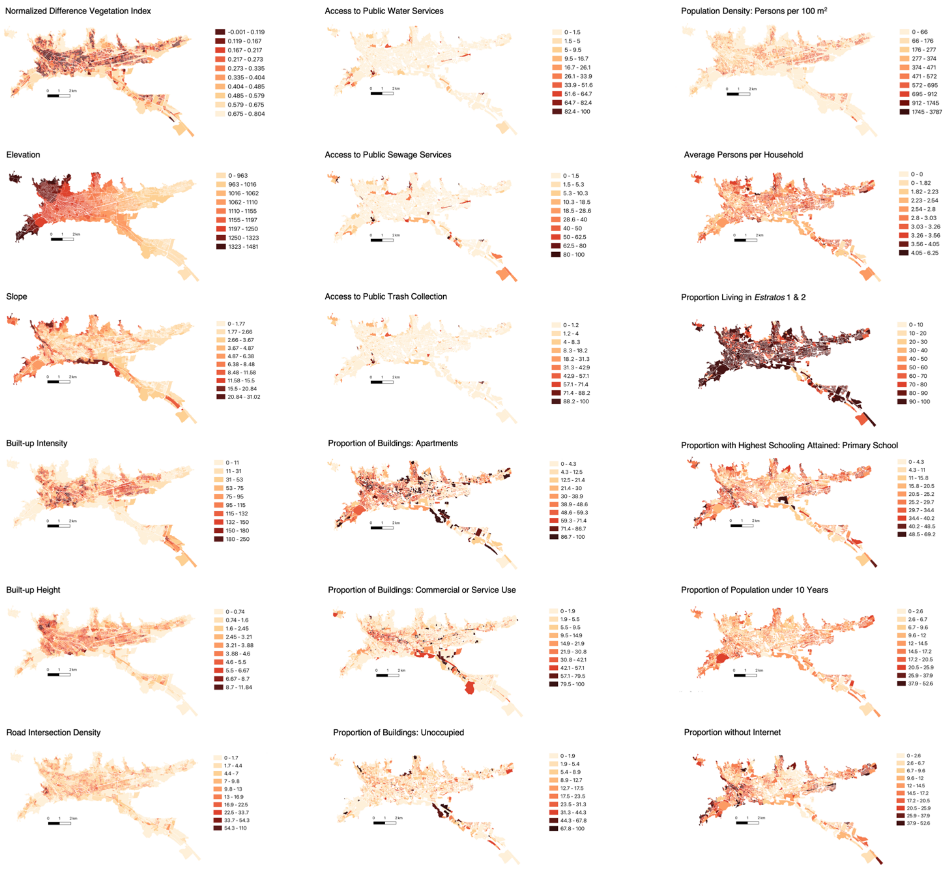


**Figure S.4 Maps of socio-environmental candidate variables at the census-block level**

Choropleth maps are shown with Jenks natural breaks to optimize the arrangement of values into “natural” classes.

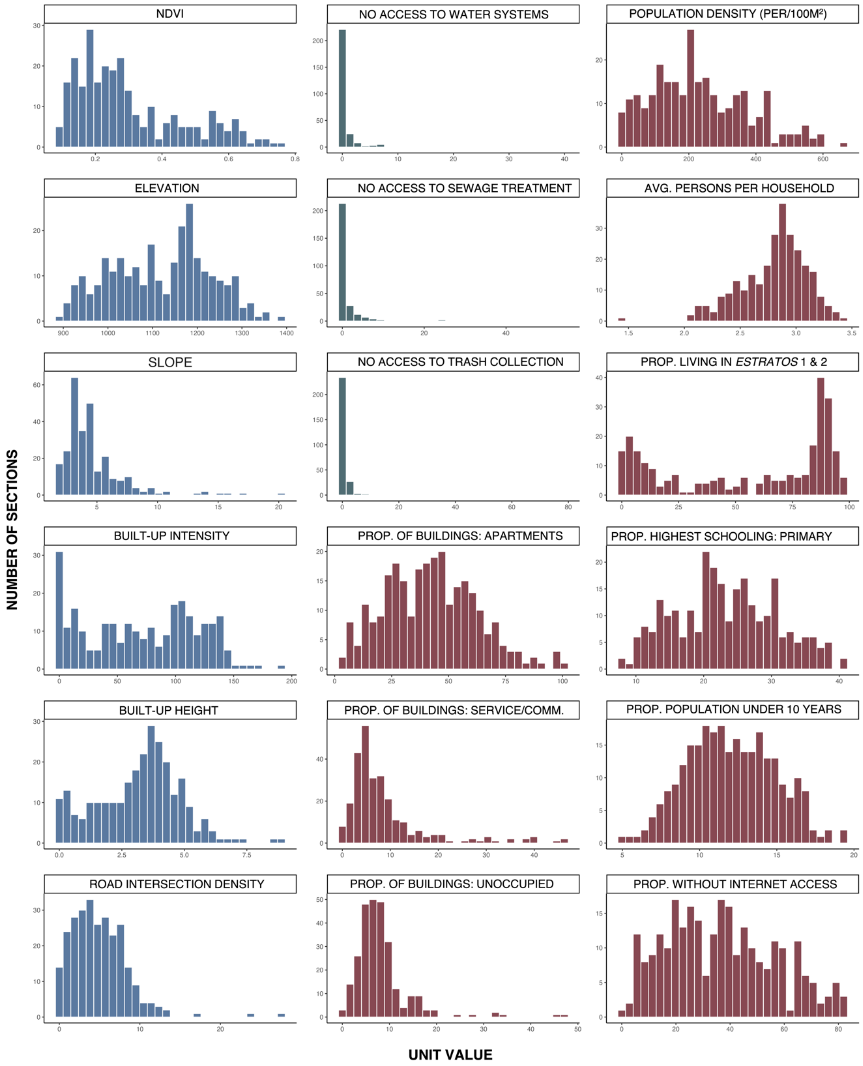


**Figure S.5 Histograms of socio-environmental covariates at the urban section level**

Covariates shown in blue represent biophysical properties; covariates in green represent institutional properties; and those in red represent socio-economic properties.

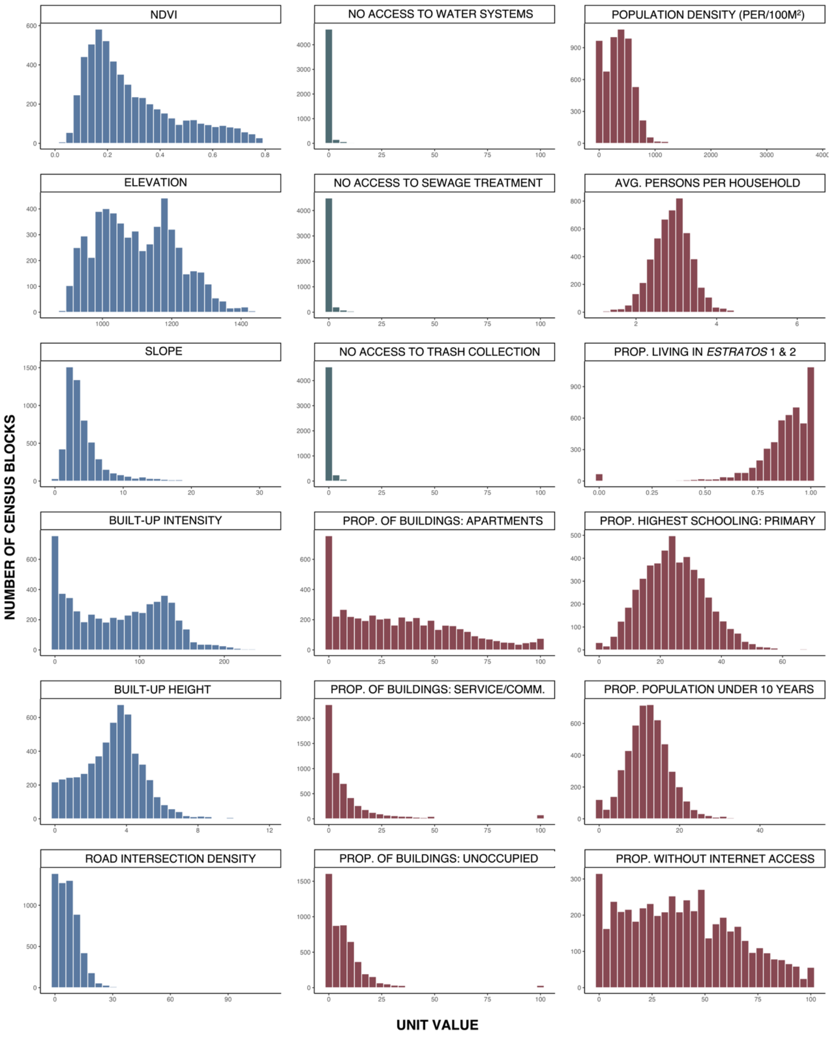


**Figure S.6 Histograms of socio-environmental covariates at the census block level**

Covariates shown in blue represent biophysical properties; covariates in green represent institutional properties; and those in red represent socio-economic properties.

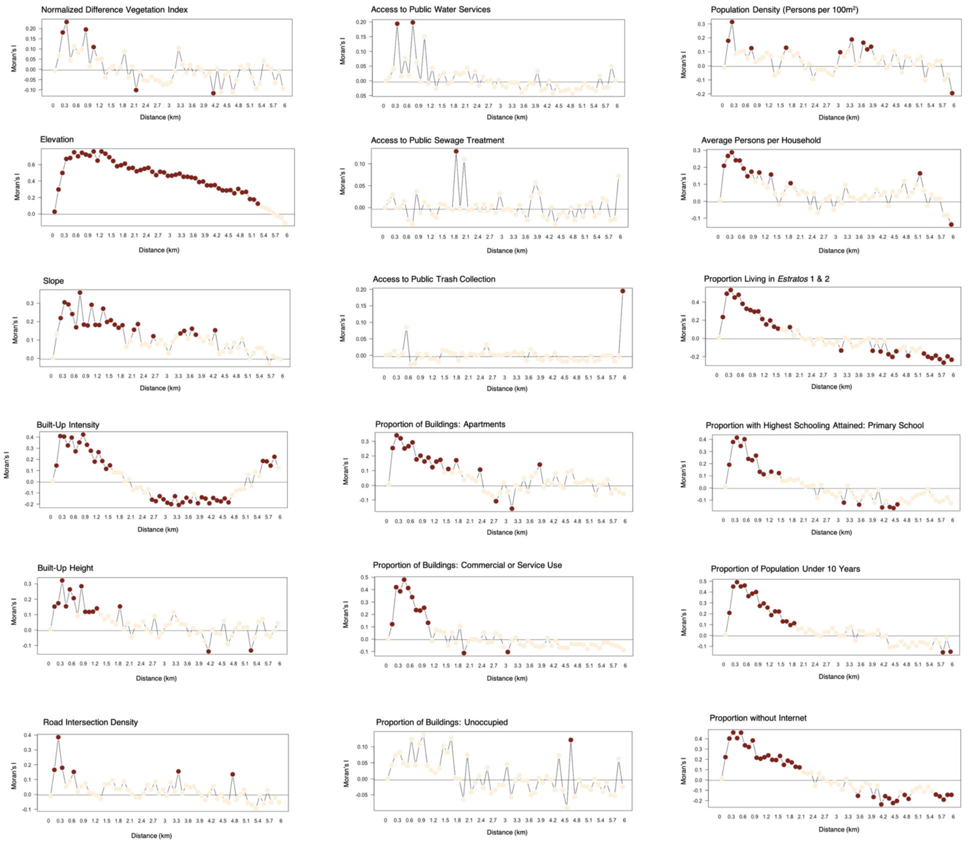


**Figure S.7 Moran's *I* spatial correlogram for covariates at the urban section level**

Red circles indicate significant spatial autocorrelation; beige circles indicate non-significant values. Significance of *I* value for each lag distance was determined with a Monte Carlo randomization test (*a* =0.05 with a progressive Bonferroni correction).

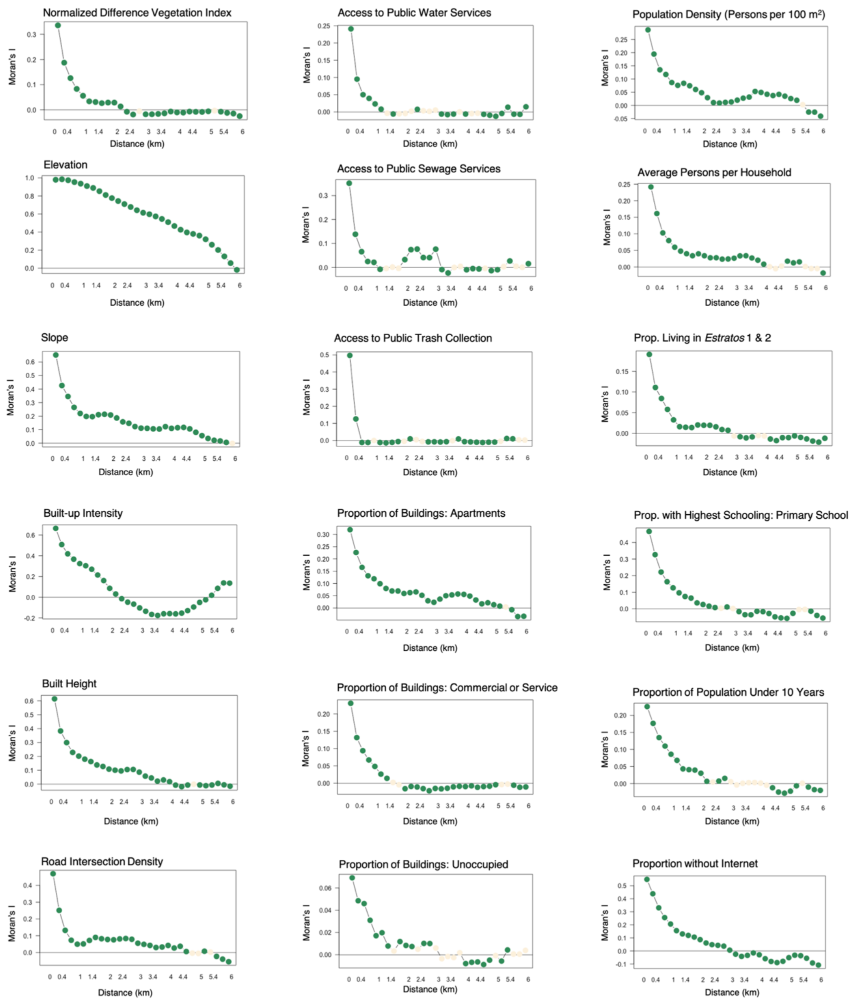


**Figure S.8 Moran's *I* spatial correlogram for covariates at the census block level**

Green circles indicate significant spatial autocorrelation; beige circles indicate non-significant values. Significance of *I* value for each lag distance was determined with a Monte Carlo randomization test (*a* =0.05 with a progressive Bonferroni correction).

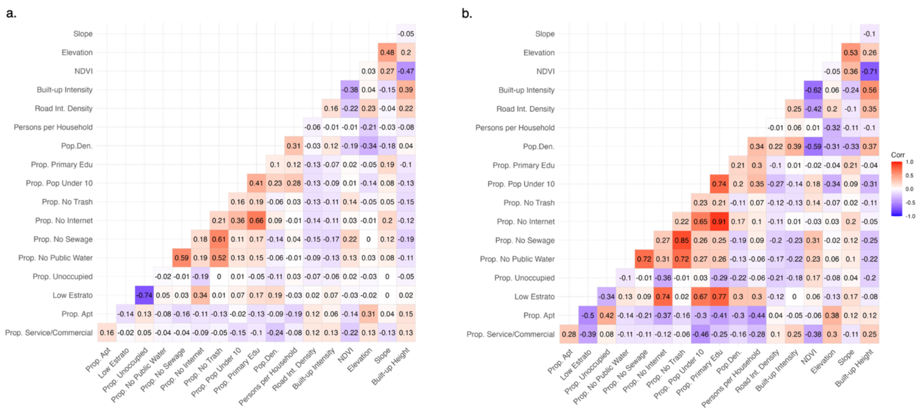


**Figure S.9 Correlation matrix of candidate variables at census block and urban section levels**

**a.** Low *estrato* neighborhoods were highly negatively correlated with the proportion of buildings unoccupied; while institutional variables including access to public water, sewage, and trash systems were highly positively correlated. Measures of the biophysical landscape, including vegetation indices (NDVI) and built-up intensity were moderately correlated. **b.** Urban section-level values were higher than the census-block level. Low *estrato* neighborhoods were highly negatively correlated with socio-economic measures, including the proportion of residences without internet access and the proportion of the population under 10 years of age. Institutional variables including access to public water, sewage, and trash systems were highly positively correlated. Measures of the biophysical landscape, including vegetation indices (NDVI) and built-up intensity were also highly correlated.
